# Serum Metabolites Associated with Depression and Anxiety in the Hispanic Community Health Study/Study of Latinos

**DOI:** 10.1101/2025.10.15.25338100

**Authors:** Xinye Qiu, Yu Zhang, Yogesh Purushotham, Qibin Qi, Bing Yu, Jan Bressler, Krista M. Perreira, Humberto Parada, Martha L. Daviglus, Alejandra Cid Vega, Carmen R. Isasi, Linda C. Gallo, Tamar Sofer

**Author notes:** **Corresponding author:** Xinye Qiu, PhD, Center for Life Sciences Floor 9, Beth Israel Deaconess Medical Center, 3 Blackfan Street, Boston, MA, USA, 02115.

## Abstract

Omics signatures for psychiatric conditions have been under-investigated. This study aimed to identify serum metabolites associated with symptoms of depression or anxiety in Hispanic/Latino adults and to develop and assess metabolite risk scores (MRSs) of these phenotypes. Using data from the Hispanic Community Health Study/Study of Latinos (HCHS/SOL), we conducted a metabolome-wide association study of 768 metabolites with depression and anxiety symptoms. We used two separate assay batches measured from baseline samples collected between June 2008 and July 2011 for discovery (batch 1: n=4,002 adults) and replication (batch 2: n=2,178 adults). We estimated the associations via survey-based generalized linear regression, and applied Least Absolute Shrinkage and Selection Operator (LASSO) regression for metabolite selection and for obtaining metabolite risk score (MRS) weights. In analyses adjusted for age, sex, field center and Hispanic/Latino background, we identified five metabolites with replicated associations with depression symptoms and six metabolites with anxiety symptoms. Key pathways related to the identified metabolites included arginine and proline, vitamin A, phospholipid, fatty acid and tyrosine metabolism. MRSs were associated with higher depression symptom score (1.07 points, 95% CI: 0.67 – 1.46, per 1 SD increase in depression MRS) and anxiety symptom score (1.05 points, 95% CI: 0.67 – 1.43, per 1 SD increase in anxiety MRS) in the replication dataset. Our study supports the associations between serum metabolites and symptoms of depression and anxiety in Hispanic/Latino adults. The MRSs developed in this study may facilitate earlier and more objective screening of individuals for depression and anxiety apart from traditional risk factors.

## 1. Introduction

Hispanic/Latino populations, compared to non-Hispanic White populations, are exposed to a higher level of adversity from stressful life events and every day stressors, and lower individual and environmental socioeconomic status.(**1, 2, 3**) These environmental stressors coupled with other behavioral risk factors can lead to a higher risk of developing chronic illness and lower quality of life, including cardiometabolic disorders and adverse neuropsychological outcomes.(**4, 5, 6**). Researchers have found that the age and sex-adjusted lifetime prevalence rates for any depressive and any anxiety disorders were about 15.4% and 15.7%, respectively in the National Latino and Asian American Study data collected from May 2002 to November 2003. In addition, Puerto Ricans had the highest prevalences (depressive disorder: 19.6%, anxiety disorder: 21.7%) as compared to the other Latino groups.(**7**) A more recent study from the Hispanic Community Health Study/Study of Latinos (HCHS/SOL) reported that the baseline (2008–2011) prevalence of elevated depression symptoms varied across Hispanic/Latino background groups, from 22.3% (Mexican) to 38.0% (Puerto Rican), indicating high and unequally distributed burden as compared to the general population.(**8**)

Past research that characterizes psychiatric conditions in terms of their genetic basis or molecular profiles including microRNAs, lipidomics, proteomics, and metabolomics have been primarily conducted in non-Hispanic White populations.(**9, 10, 11, 12, 13**) In particular, metabolomic signatures for psychiatric health have yet to be investigated in Hispanic/Latino populations. Metabolomics, the study of metabolites or small molecules produced or used during metabolism, can provide molecular biomarkers useful for disease prediction, diagnosis, prognosis, and subtyping.(**14**) Individuals with mental illnesses such as depression and anxiety disorders were observed to have differential metabolic profiles compared to healthy individuals.(**15, 16, 17, 18, 19**) In a pooled analysis of 230 metabolic markers from 10,145 control participants and 5,283 persons with depression primarily from the Netherlands, researchers found that there was a clear distinctive profile of circulating lipid metabolites associated with depression, including higher levels of apolipoprotein B, very-low-density lipoprotein cholesterol and triglycerides, and lower levels of high-density lipoprotein cholesterol, acetate, and apolipoprotein A1.(**15**) Similarly, a population-based longitudinal study of 211,200 participants from the Apolipoprotein-Related Mortality Risk cohort (mainly from the Stockholm region in Sweden) revealed that high levels of glucose and triglycerides and low levels of high-density lipoprotein were associated with increased incidence of depression, anxiety, and stress-related disorders.(**20**) Interestingly, results were similar for male and female participants. In a review study of metabolomic biomarkers in anxiety disorders, some of the most consistent findings were with metabolites related to oxidative stress, inflammatory processes, lipid and energy metabolism, glutamine metabolism, and neurotransmission.(**17**) In the HCHS/SOL Youth study of Hispanic/Latino children, the association between psychological distress and inflammation was suggested to be mediated by adiposity.(**21**) These past studies suggest there is a close relationship between lipid profiles and psychological conditions, and a metabolomic signature overlapping with that of cardiometabolic conditions.(**16**) However, while the relevant evidence is being established, existing studies are mainly based on observational cohorts of White participants with limited investigation on potential modification by sex. Metabolomics profiles for mental health phenotypes in Hispanic/Latino individuals remain limited and may not align well with the findings from White population samples given the metabolic diversity in human populations and correlation with genetic and ancestral geographic distances.(**22**)

In this study, we aimed to identify serum metabolites associated with symptoms of depression or anxiety in Hispanic/Latino adults, and to develop depression and anxiety associated metabolite risk scores (MRSs). An MRS combines the levels of multiple metabolites into a single score, potentially improving risk prediction beyond traditional risk factors, and leading to earlier and more targeted interventions.(**23, 24**) Current screening practices for depression and anxiety rely largely on subjective self-reported symptoms and may be limited to those with mental healthcare coverage. We hypothesized that certain metabolites would be associated with either higher or lower depression and anxiety symptoms. In addition, we hypothesized that metabolomic signatures may differ by sex because there are differential biological processes influenced by the sex chromosomes and reproductive organs, along with differential external environmental exposures and sensitivity to various sociodemographic, acculturation, and psychological factors.(**25**) Finally, we hypothesized that MRS developed from the current analysis would have high correlation with the mental health scores they are based on, and will have the potential to aid objective clinical screening of individuals with high risk for depression and anxiety.

## 2. Methods

### 2.1 Study population

We included participants from the HCHS/SOL, a community-based prospective cohort study of 16,415 self-identified Hispanic/Latino adults enrolled from June 2008 to July 2011 (baseline). Individuals from four field centers (Chicago, IL; Miami, FL; Bronx, NY; San Diego, CA) were recruited through a stratified multi-stage area probability sampling design process.(**26**)

Participants underwent physical exams, provided blood samples, and completed multiple questionnaires which included mental health questionnaires. Participants self-identified as either Mexican, Puerto Rican, Cuban, Central American, Dominican, South American, or as more than one or another heritage. Additional design details are provided in a previous publication.(**27**) In this study, we included 6,180 participants with available metabolomics data. Participants were grouped into two “batches” based on the analysis of samples in separate efforts for metabolite quantification. There were 4,002 from “batch 1” (the first quantification of metabolites), and 2,178 separate individuals from “batch 2” (a second quantification of metabolites). The HCHS/SOL study was approved by the institutional review boards (IRBs) at each field center, where all participants provided written informed consent, and by the non-biomedical IRB at the University of North Carolina at Chapel Hill, to the HCHS/SOL Data Coordinating Center. All methods and analyses of HCHS/SOL participants’ materials and data were carried out in accordance with human subject research guidelines and regulations.

### 2.2 Assessment of depression and anxiety

Our primary outcomes of interest were depression and anxiety symptoms. At baseline, depression symptoms were assessed using the Anderson version of the 10-item Center for Epidemiological Studies of Depression Scale (CES-D-10).(**28**) The CES-D-10 (score of 0-30) captured core self-reported symptoms of depression in the past week including depressed mood and behavioral symptoms (e.g., felt depressed; felt lonely; could not get going). The CES-D-10 has good predictive accuracy (k = 0.97 relative to the 20-item CES-D.(**28**)), and has excellent internal consistency (Cronbach’s alphas = 0.80–0.86) and test-retest reliability (Spearman’s rank correlation coefficients, r_s_ = 0.41–0.70).(**29**) The CES-D-10 total score was constructed by summing responses to each item which ranged from 0 = rarely or none of the time (< 1 day) to 3 = all of the time (5–7 days)). Higher total scores indicated higher levels of depression symptoms. Anxiety symptoms were measured using the abbreviated 10-item Spielberger State-Trait Anxiety Inventory (STAI-10, State Anxiety Subscale) which has excellent internal consistency (Cronbach’s alpha = 0.93) in the full HCHS/SOL sample.(**29**) The STAI-10 scale is a short form version of the original 20 item STAI which is a well-established measure of trait anxiety symptoms in population-based studies and correlates well with the other established anxiety measures (Taylor Manifest Anxiety Scale, and Cattell and Scheier’s Anxiety Scale Questionnaire).(**30**) For each questionnaire item, participants were asked to indicate how often (in general) (0 = almost never; 1 = sometimes; 2 = often; 3 = almost always) they felt worried, inadequate, nervous, or experienced other symptoms. Symptom scores were summed across the 10 different items following the scoring manual.(**31**) Later, 1 was added to all responses (converting from 0-3 to 1-4 scale to match the standardized instrument scoring). Higher total scores (ranging from 10 to 40) indicated higher levels of anxiety symptoms.

### 2.3 Metabolomics measurement and processing

Fasting blood samples were collected, processed, and stored at −70 °C until laboratory analysis. A total of 4,002 individuals were selected at random (out of the set of participants who had whole genome sequencing data, itself a random subset of the study population) for metabolomics assessment in batch 1. Later, a second set of individuals were profiled in batch 2 based on the following criteria (**32**): 1) including 50 samples for quality control analyses between the first and second batch; 2) adding individuals who did not have metabolomics measured in batch 1, and including a subset of individuals who participated in the ECHO-SOL ancillary study (**33**) with an average age older than the entire cohort (at least 45 years old at the exam); 3) sampling at random from individuals with the most declined estimated glomerular filtration rate (eGFR) measures between baseline and a second clinic visit (visit 2)Individuals included in both batches were removed from the analysis in batch 2 (resulting in a final sample size of 2,178 individuals) so that the two batch samples were independent of each other. The metabolomic profiling was conducted at Metabolon (Durham, NC) using the Discovery HD4 platform in 2017 for batch 1 and in 2021 for batch 2. Metabolites were quantified with an untargeted, liquid chromatography-mass spectrometry (LC–MS)-based quantification protocol.(**34**) Metabolites have sporadic missingness values. We used only metabolites with ≤25% missingness in both batches (768 metabolites). Missingness in the metabolites included in the analysis was imputed using half the minimum observed value of the metabolite in the batch-specific sample under the assumption that metabolites were not observed due to being below the technical detection limit. Before modeling, we further applied rank-normalization to each included metabolite in each batch separately.

### 2.4 Statistical analyses

Figure 1 provides an overview of the analytical steps involved in the analyses.

**Figure 1.**
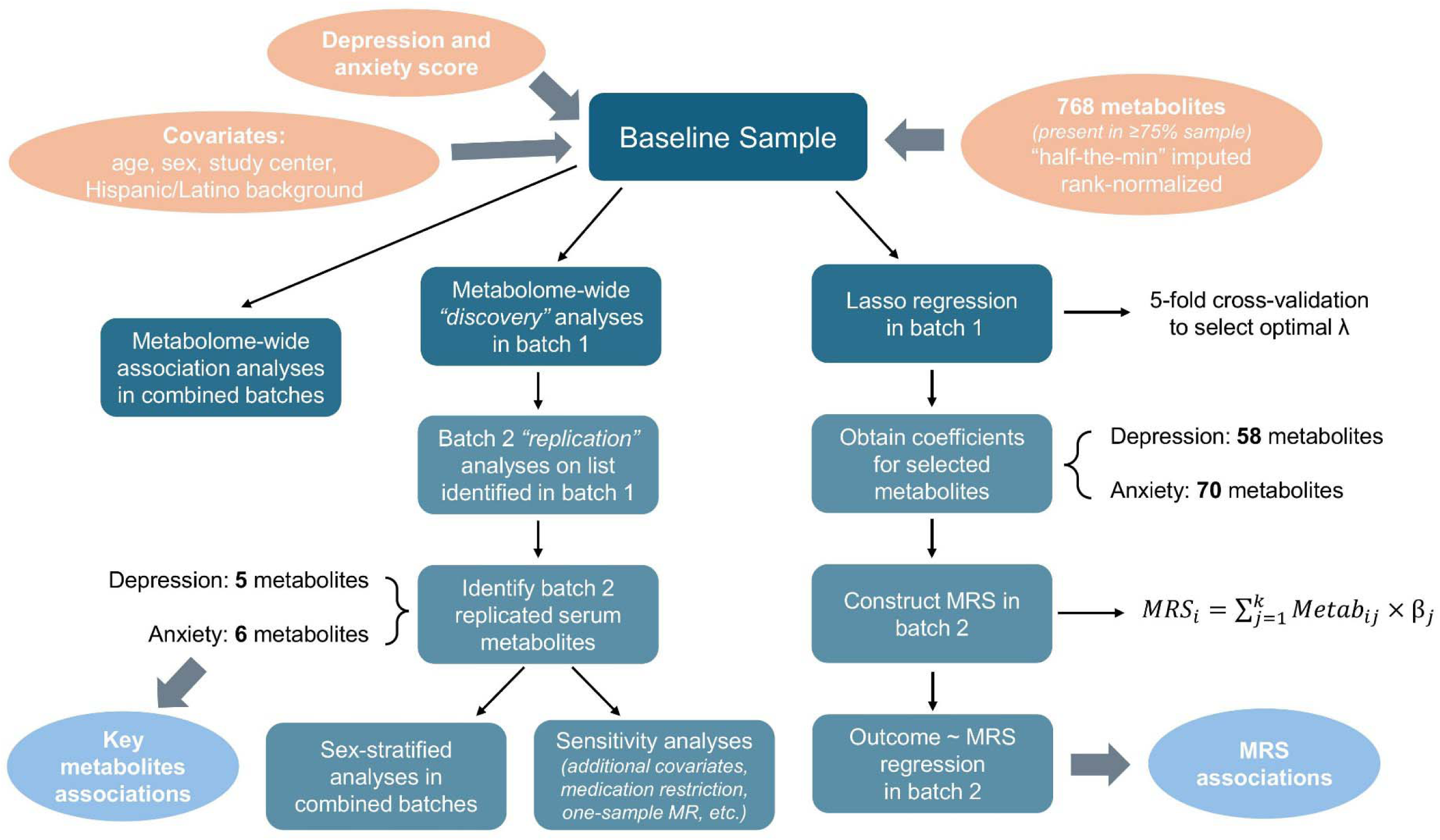
Flow Chart of Overall Analytical Approach.

#### 2.4.1 Discovery and replication analyses

We applied an analytic approach where we first performed metabolome-wide association analyses among batch 1 participants to identify metabolites associated with either depression or anxiety symptoms (discovery analysis). Following that, we conducted a replication analysis among batch 2 participants, estimating the associations of metabolites identified in the discovery analysis to assess replicated metabolite associations. We applied this approach because replication is a gold standard in large omics studies. We annotated the associated metabolic pathways for each replicated metabolite association using the information provided by the Metabolon platform.

#### 2.4.2 Covariates adjustment

We conducted three sets of covariate adjustments. The primary model (Model I) adjusted for age at baseline (continuous: in years), sex (categorical: female, male), field center (categorical: Bronx, Chicago, Miami and San Diego) and Hispanic/Latino background (categorical: Dominican, Central American, Cuban, Mexican, Puerto Rican, South American, more than one heritage). For Model II, we additionally adjusted for annual household income (6-level categorical: missing, less than $10,000, $10,001-$20,000, $20,001-$40,000, $40,001-$75,000, more than $75,000) and education (3-level categorical: <12 years, =12 years, >12 years), in addition to Model I covariates. Model III included Model II covariates and additionally adjusted for health behaviors, including physical activity level (continuous: total metabolic equivalent task in minutes per day), diet score (continuous: the Alternative Healthy Eating Index 2010, AHEI-2010), smoking (3-level categorical: never, former and current smoker) and alcohol consumption status (3-level categorical: never, former and current alcohol drinker).

#### 2.4.3 Stratified analyses by sex

We conducted sex-stratified analyses adjusted for Model I covariates that combined batches (batch 1 plus batch 2) to identify potential sex differences in metabolite associations. Here, combined batches were used to increase the power of analyses, as sex-specific analyses have limited sample sizes.

#### 2.4.4 LASSO-based metabolite risk score (MRS)

To develop a MRS for depression and anxiety symptoms, we applied Least Absolute Shrinkage and Selection Operator (LASSO) regression to select key metabolites and obtain their coefficients for each outcome in batch 1. First, a 5-fold cross-validation was applied to select the optimal tuning parameter value. LASSO was applied using the selected Lambda value over the complete batch 1 dataset. Next, coefficients from the LASSO regression for selected metabolites were extracted and used as weights in a weighted index, constructed in batch 2 samples by multiplying the corresponding, rank-normalized, metabolite levels by their weights, and summing. After obtaining the weighted index (i.e. the MRS) in batch 2, we z-scored the MRSs and estimated their associations with each mental health scale (as appropriate) in a survey-weighted linear regression adjusted for Model I covariates.

#### 2.4.5 Sensitivity analyses

We conducted several sensitivity analyses (adjusted for Model I covariates) to test the robustness of our primary findings. First, we restricted the analytic sample to those who did not use antidepressant or anti-anxiety medication, since medication use may influence the level of metabolites and interfere with the level of psychiatric symptoms. Second, we conducted similar metabolome-wide-association analyses in a combined batch dataset to increase statistical power of analyses. Third, we performed longitudinal, repeated measures analyses utilizing both visits (visit 1 and 2) and combining both batches via a mixed effects model adjusting for Model I covariates with a random intercept of each participant ID and each household ID to increase the power of analysis and account for within cluster correlations when having repeated measures. Note that this is a different approach from the survey-based analysis of a single-visit analysis, which accounts for participant clustering in a different manner. The mixed effects model was only conducted for depression symptom score (CES-D-10) since the same anxiety symptom score (STAI-10) is not available at visit 2. Last, we applied one-sample Mendelian randomization (MR) analysis. (**35**) The MR analysis was only carried out for four metabolites (3-amino-2-piperidone, hydroxy-CMPF*, carotene-diol (1) and choline) with available genome-wide association study (GWAS) summary statistics from the NETSIM Finnish population study.(**36**) We used the GWAS summary statistics for a given metabolite to construct a polygenic score as follows. Single nucleotide polymorphisms (SNPs) were filtered using a significance p value threshold of 5×10^-8^ and followed by clumping based on the 1000 genome project European reference panel (linkage disequilibrium R^2^ =0.01). In the HCHS/SOL datasets, the effect alleles were summed with weights being the estimated effect sizes from the GWAS, resulting in a polygenic score for the metabolite. Next, for each of the four metabolites, we used the ‘svyivreg’ function from the ‘survey’ package(**37**) to estimate if there is a potential causal effect of the observed metabolite on depression or anxiety symptom score using the polygenic score as an instrumental variable. All statistical models for primary and secondary analyses (except for LASSO and mixed effects model) were survey-based generalized linear models (achieved by the ‘svgglm’ function from the ‘survey’ package) incorporating sampling weights at baseline visit. R version 4.2.2 was used. Statistical significance was achieved when a two-sided false discovery rate (FDR)-corrected(**38**) p value was less than 0.05.

## 3. Results

### 3.1 Participants characteristics

Participants included in this study sample had an average age of 48.3 years at baseline (SD = 13.2), 60% were female, and 34% were of Mexican background. The majority of the participants were overweight (BMI ≥25 & <30 kg/m2, 37%) or obese (BMI ≥30 kg/m2, 43%), currently drinking alcohol (47%) and had never smoked (59%). Participant characteristics differed between the two metabolomics batches, as expected according to their selection criteria, including average age, proportions of females, Hispanic/Latino background, BMI distribution, smoking, and alcohol intake status proportions. Unweighted detailed summary characteristics for participants included in batch 1, batch 2 and combined batches can be found in **Table 1**.

**Table 1.** Participants Characteristics at Baseline by Batch Sample of Metabolites Profiling.

| **Group** | **Batch Combined** | **Batch 1** | **Batch 2** |
| --- | --- | --- | --- |
| Sample size, N | 6,180 | 4,002 | 2,178 |
| Age at baseline, yrs. (mean (SD)) | 48.28 (13.17) | 45.89 (13.80) | 52.68 (10.59) |
| Sex (n (%)) |  |  |  |
| Female | 3,704 (59.9) | 2,297 (57.4) | 1,407 (64.6) |
| Male | 2,476 (40.1) | 1,705 (42.6) | 771 (35.4) |
| Hispanic/Latino background (n (%)) |  |  |  |
| Dominican | 672 (10.9) | 392 (9.8) | 280 (12.9) |
| Central American | 638 (10.3) | 407 (10.2) | 231 (10.6) |
| Cuban | 1,082 (17.5) | 677 (16.9) | 405 (18.6) |
| Mexican | 2,105 (34.1) | 1,441 (36.0) | 664 (30.5) |
| Puerto-Rican | 1,104 (17.9) | 716 (17.9) | 388 (17.8) |
| South American | 409 (6.6) | 234 (5.8) | 175 (8.0) |
| More than one or other heritage | 155 (2.5) | 126 (3.1) | 29 (1.3) |
| Education category (n (%)) |  |  |  |
| < 12 years | 2,282 (37.0) | 1,444 (36.1) | 838 (38.6) |
| 12 years | 1,541 (25.0) | 1,025 (25.7) | 516 (23.8) |
| >12 years | 2,343 (38.0) | 1,527 (38.2) | 816 (37.6) |
| BMI category (n (%)) |  |  |  |
| Underweight (BMI < 18.5 kg/m<sup>2</sup>) | 39 (0.6) | 33 (0.8) | 6 (0.3) |
| Healthy Weight (BMI ≥18.5 & <25 kg/m<sup>2</sup>) | 1,159 (18.8) | 798 (20.0) | 361 (16.6) |
| Overweight (BMI ≥25 & <30 kg/m<sup>2</sup>) | 2,305 (37.4) | 1,458 (36.6) | 847 (39.0) |
| Obese (BMI ≥ 30 kg/m<sup>2</sup>) | 2,658 (43.1) | 1,699 (42.6) | 959 (44.1) |
| Alcohol intake category (n (%)) |  |  |  |
| Never | 1,275 (20.7) | 739 (18.5) | 536 (24.7) |
| Former | 2,005 (32.5) | 1,289 (32.2) | 716 (33.0) |
| Current | 2,891 (46.8) | 1,973 (49.3) | 918 (42.3) |
| Smoking category (n (%)) |  |  |  |
| Never | 3,612 (58.6) | 2,340 (58.5) | 1,272 (58.6) |
| Former | 1,292 (20.9) | 796 (19.9) | 496 (22.9) |
| Current | 1,264 (20.5) | 863 (21.6) | 401 (18.5) |

### 3.2 Findings from primary metabolome-wide association analysis

**Table 2** shows the estimated effect sizes and 95% confidence intervals (CIs) of replicated metabolite associations with depression and anxiety symptom scores from Model I, accounting for age, sex, field center, and Hispanic/Latino background. Associations are shown per 1 SD increase in rank-normalized metabolite levels. Among metabolites replicated in batch 2, we identified five metabolites with higher concentrations associated with levels of depression symptoms. Tyramine O-sulfate, choline, and the unknown metabolite X-24337 were associated with higher depression symptom scores, whereas hydroxy-CMPF and beta-cryptoxanthin were associated with lower depression symptom scores. Six metabolites had replicated associations with anxiety symptoms: tyramine O-sulfate, 3-amino-2-piperidone, and X-24337 were associated with higher anxiety symptom scores, while carotene diol (1), hydroxy-CMPF, and X-11315 were associated with lower anxiety symptom scores. These observed associations weakened upon adjustment for additional covariates in Models II and III. However, several remained significantly associated with the outcome symptom scores for both batches under Model II and III after FDR correction. For depression symptoms, all identified metabolites remained associated in both batches under Model II. With Model III adjustment, beta-cryptoxanthin and choline remained associated with depression symptoms (beta-cryptoxanthin: batch 1: -0.34, 95% CI: -0.60 to -0.07; batch 2: -0.52, 95% CI: -0.96 to -0.09; choline: batch 1: 0.36, 95% CI: 0.13 to 0.59; batch 2: 0.45, 95% CI: 0.07 to 0.84). Levels of tyramine O-sulfate, carotene diol (1), 3-amino-2-piperidone and unknown X-11315 remained associated with anxiety symptom scores while additionally accounting for personal income and education in both batches under Model II. After additionally accounting for health behavioral factors under Model III adjustment, their associations were not replicated in both batches. However, we still observed associations with similar pattern and similar effect magnitude as compared with Model I findings for all identified metabolites. We observed low to moderate correlations (Pearson correlation r: - 0.36 ∼ 0.54) among the identified metabolites associated with depression or anxiety symptoms (**Figure S1** in the Supplement).

**Table 2.**
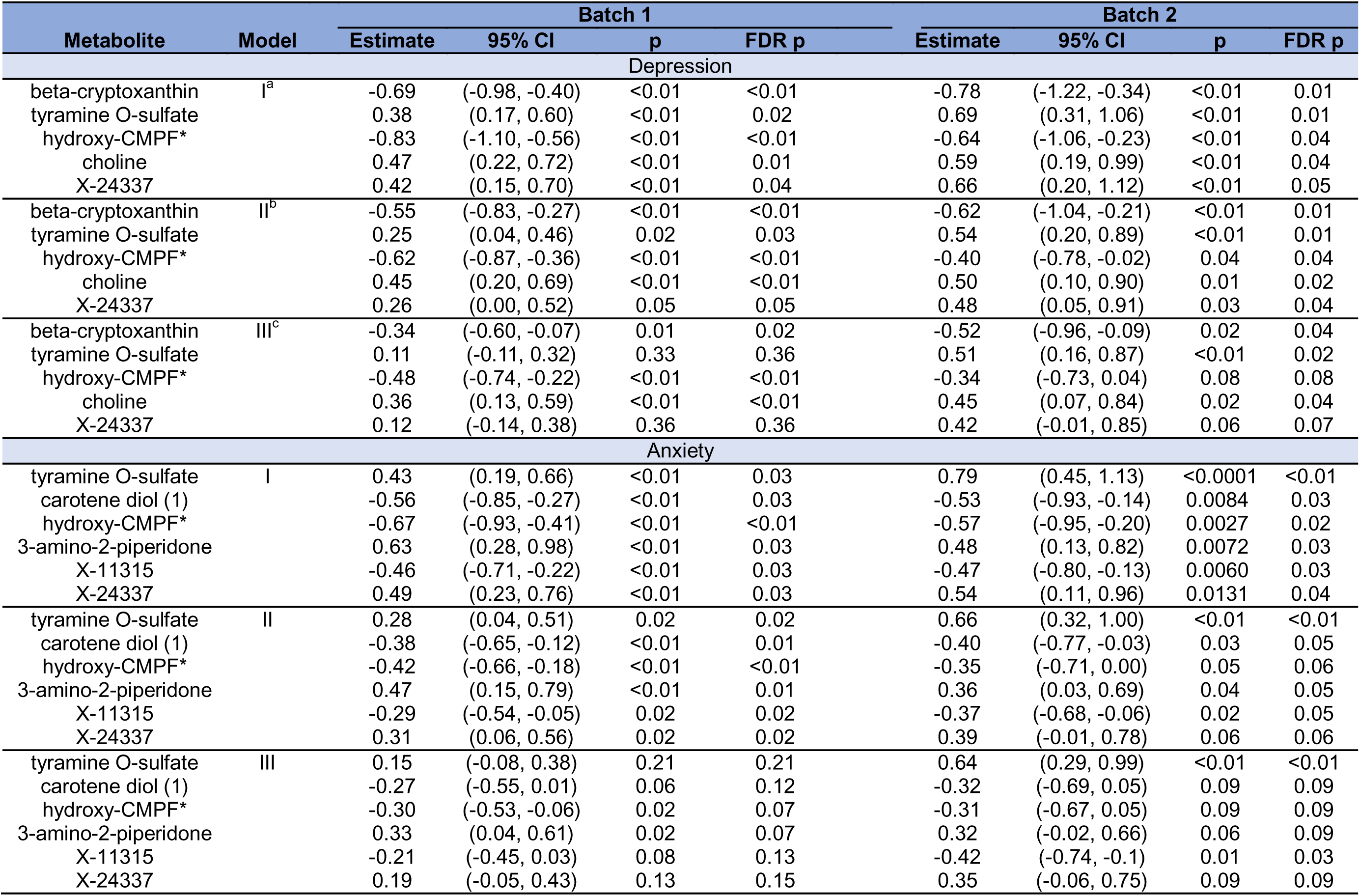
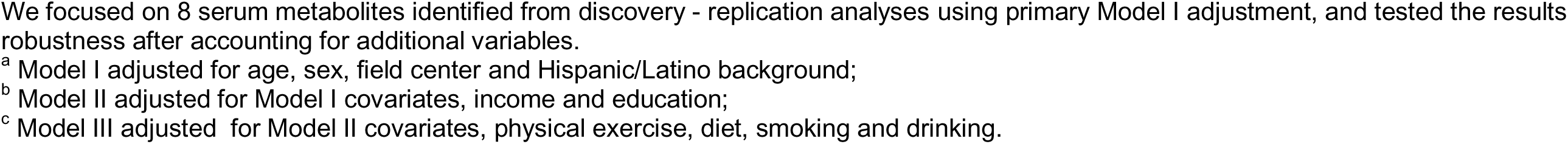
Association Results for Identified Metabolites Associated with Depression and Anxiety Symptoms.

| Metabolite | Model | Batch 1 |  |  |  | Batch 2 |  |  |  |
| --- | --- | --- | --- | --- | --- | --- | --- | --- | --- |
|  |  | Estimate | 95% CI | p | FDR p | Estimate | 95% CI | p | FDR p |
| Depression |  |  |  |  |  |  |  |  |  |
| beta-cryptoxanthin | I<sup>a</sup> | -0.69 | (-0.98, -0.40) | <0.01 | <0.01 | -0.78 | (-1.22, -0.34) | <0.01 | 0.01 |
| tyramine O-sulfate |  | 0.38 | (0.17, 0.60) | <0.01 | 0.02 | 0.69 | (0.31, 1.06) | <0.01 | 0.01 |
| hydroxy-CMPF* |  | -0.83 | (-1.10, -0.56) | <0.01 | <0.01 | -0.64 | (-1.06, -0.23) | <0.01 | 0.04 |
| choline |  | 0.47 | (0.22, 0.72) | <0.01 | 0.01 | 0.59 | (0.19, 0.99) | <0.01 | 0.04 |
| X-24337 |  | 0.42 | (0.15, 0.70) | <0.01 | 0.04 | 0.66 | (0.20, 1.12) | <0.01 | 0.05 |
| beta-cryptoxanthin | II<sup>b</sup> | -0.55 | (-0.83, -0.27) | <0.01 | <0.01 | -0.62 | (-1.04, -0.21) | <0.01 | 0.01 |
| tyramine O-sulfate |  | 0.25 | (0.04, 0.46) | 0.02 | 0.03 | 0.54 | (0.20, 0.89) | <0.01 | 0.01 |
| hydroxy-CMPF* |  | -0.62 | (-0.87, -0.36) | <0.01 | <0.01 | -0.40 | (-0.78, -0.02) | 0.04 | 0.04 |
| choline |  | 0.45 | (0.20, 0.69) | <0.01 | <0.01 | 0.50 | (0.10, 0.90) | 0.01 | 0.02 |
| X-24337 |  | 0.26 | (0.00, 0.52) | 0.05 | 0.05 | 0.48 | (0.05, 0.91) | 0.03 | 0.04 |
| beta-cryptoxanthin | III<sup>c</sup> | -0.34 | (-0.60, -0.07) | 0.01 | 0.02 | -0.52 | (-0.96, -0.09) | 0.02 | 0.04 |
| tyramine O-sulfate |  | 0.11 | (-0.11, 0.32) | 0.33 | 0.36 | 0.51 | (0.16, 0.87) | <0.01 | 0.02 |
| hydroxy-CMPF* |  | -0.48 | (-0.74, -0.22) | <0.01 | <0.01 | -0.34 | (-0.73, 0.04) | 0.08 | 0.08 |
| choline |  | 0.36 | (0.13, 0.59) | <0.01 | <0.01 | 0.45 | (0.07, 0.84) | 0.02 | 0.04 |
| X-24337 |  | 0.12 | (-0.14, 0.38) | 0.36 | 0.36 | 0.42 | (-0.01, 0.85) | 0.06 | 0.07 |
| Anxiety |  |  |  |  |  |  |  |  |  |
| tyramine O-sulfate | I | 0.43 | (0.19, 0.66) | <0.01 | 0.03 | 0.79 | (0.45, 1.13) | <0.0001 | <0.01 |
| carotene diol (1) |  | -0.56 | (-0.85, -0.27) | <0.01 | 0.03 | -0.53 | (-0.93, -0.14) | 0.0084 | 0.03 |
| hydroxy-CMPF* |  | -0.67 | (-0.93, -0.41) | <0.01 | <0.01 | -0.57 | (-0.95, -0.20) | 0.0027 | 0.02 |
| 3-amino-2-piperidone |  | 0.63 | (0.28, 0.98) | <0.01 | 0.03 | 0.48 | (0.13, 0.82) | 0.0072 | 0.03 |
| X-11315 |  | -0.46 | (-0.71, -0.22) | <0.01 | 0.03 | -0.47 | (-0.80, -0.13) | 0.0060 | 0.03 |
| X-24337 |  | 0.49 | (0.23, 0.76) | <0.01 | 0.03 | 0.54 | (0.11, 0.96) | 0.0131 | 0.04 |
| tyramine O-sulfate | II | 0.28 | (0.04, 0.51) | 0.02 | 0.02 | 0.66 | (0.32, 1.00) | <0.01 | <0.01 |
| carotene diol (1) |  | -0.38 | (-0.65, -0.12) | <0.01 | 0.01 | -0.40 | (-0.77, -0.03) | 0.03 | 0.05 |
| hydroxy-CMPF* |  | -0.42 | (-0.66, -0.18) | <0.01 | <0.01 | -0.35 | (-0.71, 0.00) | 0.05 | 0.06 |
| 3-amino-2-piperidone |  | 0.47 | (0.15, 0.79) | <0.01 | 0.01 | 0.36 | (0.03, 0.69) | 0.04 | 0.05 |
| X-11315 |  | -0.29 | (-0.54, -0.05) | 0.02 | 0.02 | -0.37 | (-0.68, -0.06) | 0.02 | 0.05 |
| X-24337 |  | 0.31 | (0.06, 0.56) | 0.02 | 0.02 | 0.39 | (-0.01, 0.78) | 0.06 | 0.06 |
| tyramine O-sulfate | III | 0.15 | (-0.08, 0.38) | 0.21 | 0.21 | 0.64 | (0.29, 0.99) | <0.01 | <0.01 |
| carotene diol (1) |  | -0.27 | (-0.55, 0.01) | 0.06 | 0.12 | -0.32 | (-0.69, 0.05) | 0.09 | 0.09 |
| hydroxy-CMPF* |  | -0.30 | (-0.53, -0.06) | 0.02 | 0.07 | -0.31 | (-0.67, 0.05) | 0.09 | 0.09 |
| 3-amino-2-piperidone |  | 0.33 | (0.04, 0.61) | 0.02 | 0.07 | 0.32 | (-0.02, 0.66) | 0.06 | 0.09 |
| X-11315 |  | -0.21 | (-0.45, 0.03) | 0.08 | 0.13 | -0.42 | (-0.74, -0.1) | 0.01 | 0.03 |
| X-24337 |  | 0.19 | (-0.05, 0.43) | 0.13 | 0.15 | 0.35 | (-0.06, 0.75) | 0.09 | 0.09 |

We focused on 8 serum metabolites identified from discovery - replication analyses using primary Model I adjustment, and tested the results robustness after accounting for additional variables.
<sup>a</sup> Model I adjusted for age, sex, field center and Hispanic/Latino background;
<sup>b</sup> Model II adjusted for Model I covariates, income and education;
<sup>c</sup> Model III adjusted for Model II covariates, physical exercise, diet, smoking and drinking.

### 3.3 Sex-specific findings

Figure 2. displays sex-specific estimates of the associations between metabolites identified in the primary, sex-combined analysis and depression and anxiety symptom scores. Overall, the metabolite – depression symptoms associations were weaker in males compared to females across the 8 metabolites assessed. Particularly, serum choline level was not associated with depression symptoms among males but was harmfully associated among females. For anxiety symptoms, the sex differences were less noticeable based on the differences in effect magnitude. However, choline level was again only harmfully associated with anxiety symptoms in females but not males. **Tables S1** and **S2** in the Supplement present the full lists of metabolites associated with the two symptom scores by sex when using a batch-combined sample, and their effect estimates, 95% CIs, p values and linked super and sub-pathways. A large proportion of the identified metabolites displayed effect heterogeneity by sex with association strengths more attenuated towards the null for males (except for several metabolites in the pathways of carnitine, steroids, polyamine, phosphatidylethanolamine, phosphatidylinositol and phosphatidylcholine metabolism). Details can be viewed in the sex-stratified forest plot **Figure S2** in the Supplement.

**Figure 2.**
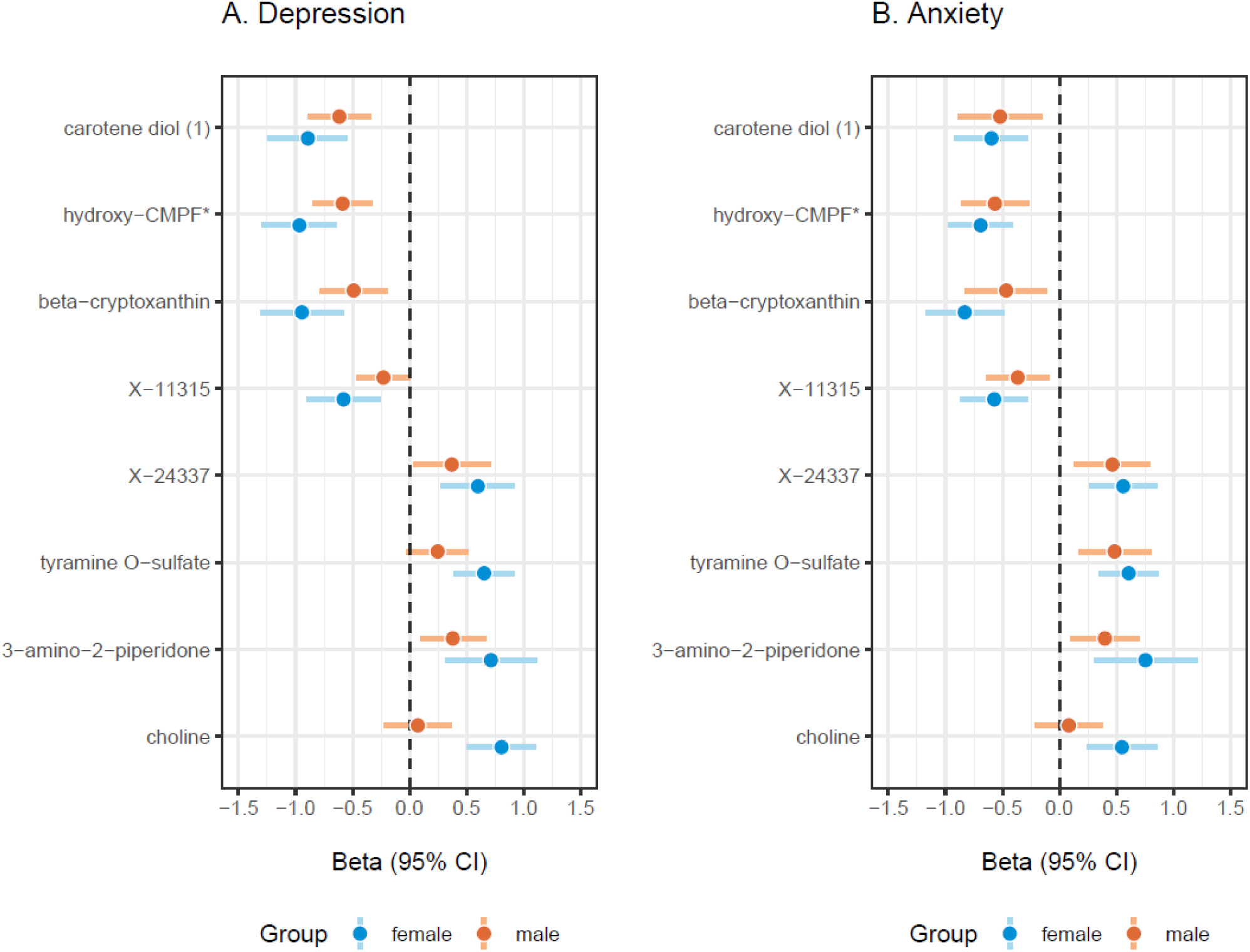
Sex Stratified Associations Between 8 Identified Serum Metabolites Associated with Depression (CES-D-10) and Anxiety (STAI-10) Symptom Scores. Beta coefficient (beta) indicates the difference of depression or anxiety score per 1 standard deviation (SD) increase in associated serum metabolites level after adjusting for age, sex, field center and Hispanic/Latino origins in batch combined sample. The color bar covers the 95% confidence interval while the middle point is the point estimate of beta. Blue bars stand for females while red bars stand for males.

### 3.4 Metabolite risk scores

Metabolome-wide LASSO regression was applied to select a set of metabolites jointly associated with depression or anxiety symptoms while accounting for the influence of all measured metabolites. A total of 58 metabolites were identified for depression symptom score and a total of 70 metabolites were identified for anxiety symptoms. Among the LASSO identified metabolite list for depression and anxiety symptoms, a total of 34 metabolites overlapped. **Table S3** in the Supplement displays the details of the unique and overlapping metabolites across the two sets of identified metabolites. The summation of selected metabolites level (in batch 2) and their beta coefficients from LASSO (in batch 1) were then z-scored to generate the metabolite risk score (MRS). Distributions of the loading coefficients are displayed in Figure 3. The constructed MRS showed significant association with each mental health indicator - a 1.07 point increase in depression (CES-D-10) score (95% CI: 0.67 to 1.46) per 1 SD increase in depression MRS (p <0.001) and a 1.05 point increase in anxiety (STAI-10) score (95% CI: 0.67 to 1.43) per 1 SD increase in anxiety MRS (p <0.001).

**Figure 3.**
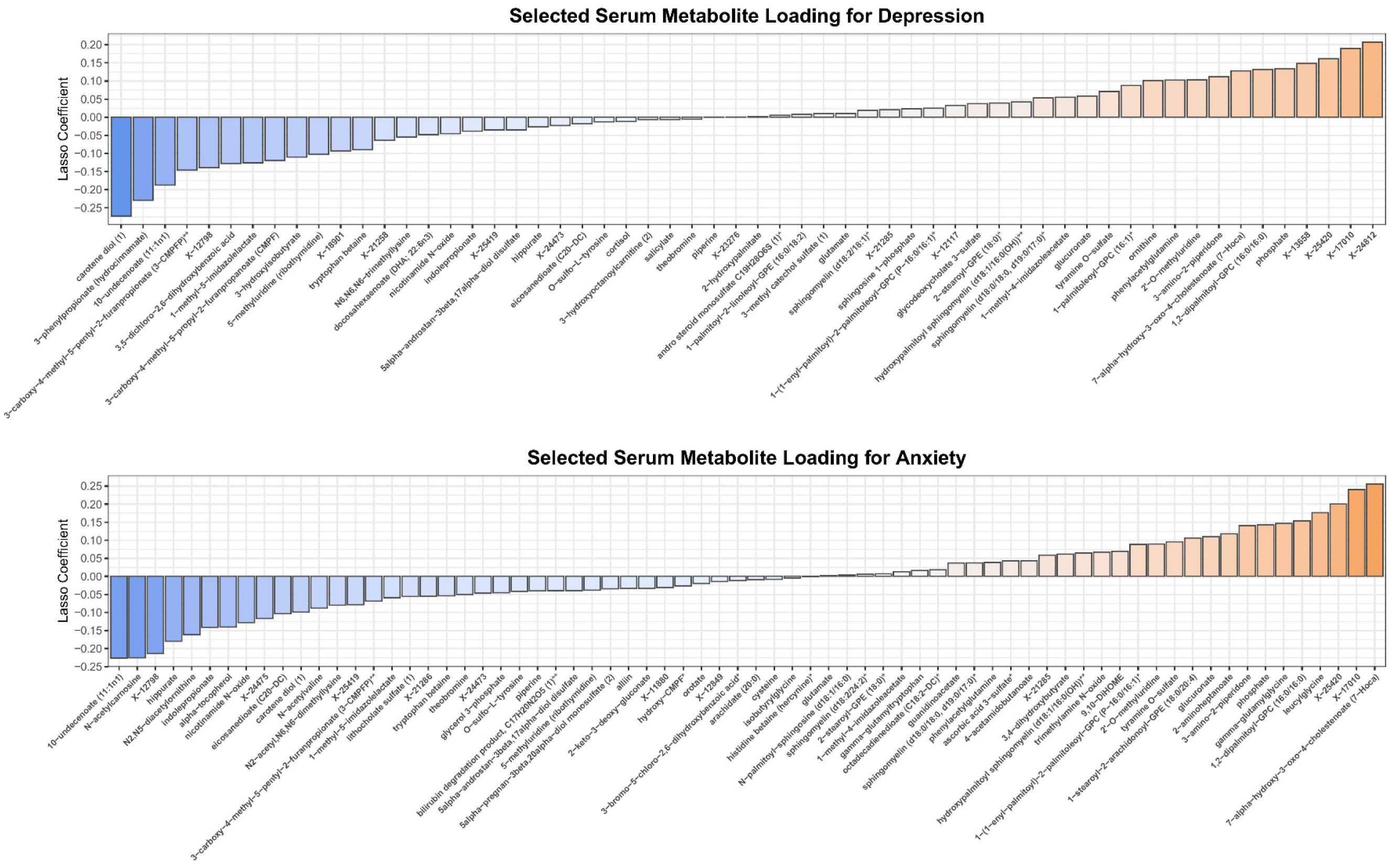
Distributions of Loading Coefficients for Selected Serum Metabolites Based on LASSO Adjusted for Age, Sex, Field Center and Hispanic/Latino Origins.

### 3.5 Identified metabolic pathways

**Table 3**. shows the specific super- and sub-pathways for the eight identified replicated metabolites. In general, super-pathways of amino acid, vitamins and lipid metabolism were represented. Specifically, two out of the eight metabolites (beta-cryptoxanthin and carotene diol (1)) were involved in vitamin A metabolism. Choline, the most robustly associated metabolite for depression symptoms, was involved in the pathway of phospholipid metabolism.

**Table 3.** Outcome Associations and Involved Metabolic Pathways for Identified Serum Metabolites.

| Metabolite | Outcome Links<sup>a</sup> | Super Pathway | Sub Pathway | HMDB |
| --- | --- | --- | --- | --- |
| 3-amino-2-piperidone | anxiety (+) | Amino Acid | Urea cycle; Arginine and Proline Metabolism | HMDB0000323 |
| beta-cryptoxanthin | depression (-) | Cofactors and Vitamins | Vitamin A Metabolism | HMDB0033844 |
| carotene diol (1) | anxiety (-) | Cofactors and Vitamins | Vitamin A Metabolism | N/A |
| choline | depression (+) | Lipid | Phospholipid Metabolism | HMDB0000097 |
| hydroxy-CMPF* | depression (-), anxiety (-) | Lipid | Fatty Acid, Dicarboxylate | N/A |
| tyramine O-sulfate | depression (+), anxiety (+) | Amino Acid | Tyrosine Metabolism | HMDB0006409 |
| X-11315 | anxiety (-) | N/A<sup>b</sup> | N/A | N/A |
| X-24337 | depression (+), anxiety (+) | N/A | N/A | N/A |

<sup>a</sup> decrease -; increase: + of depression or anxiety symptoms;
<sup>b</sup> N/A indicates missing information.

### 3.6 Sensitivity analyses

After restricting participants to those not using antidepressants or anti-anxiety medications at baseline, four of the five identified metabolites remained associated with depression symptoms under Model I adjustment. Also, five of the six identified metabolites remained associated with anxiety symptoms under Model I adjustment, supporting the robustness of the primary findings. Details can be viewed in **Table S4** in the Supplement. Batch-combined analyses, more powerful due to larger sample size, identified additional metabolites (85 total for depression and 50 for anxiety), though replication testing was not available (**Table S5** and **S6** in the Supplement).

Figure S3 in the Supplement displays the distribution of effect estimates for each identified metabolite from the batch-combined analyses. All replicated metabolite associations from the primary analyses remained associated in batch-combined modeling. Beta-cryptoxanthin, carotene diol (1) and hydroxy-CMPF* had strong protective associations with both depression and anxiety symptoms based on batch-combined analyses. In addition, 3-amino-2-piperidone is the top metabolite positively associated with both symptoms in batch-combined analysis. All five metabolites identified to be linked with depression symptoms from primary models remained associated in mixed effects models among 6,997 individuals with either metabolite measured at baseline visit or visit 2, or both visits from both batches (**Table S7** in the Supplement). MR analyses based on GWAS of plasma metabolites from the Finnish population (n = 6,136) did not support causal effects of 3-amino-2-piperidone, hydroxy-CMPF*, carotene-diol (1) or choline on depression or anxiety symptoms, despite evidence that the polygenic scores were strong instrumental variables for their respective metabolites. Details of the effect estimates, and relevant information are described in **Table S8** in the Supplement.

## 4. Discussion

In this study, we used two separate subsets of individuals from the HCHS/SOL to identify and replicate blood metabolites associations with depression and anxiety symptoms in Hispanic/Latino adults. Using a metabolome-wide association analyses approach, several blood metabolites were identified to be associated with levels of self-reported depression (beta-cryptoxanthin, choline, hydroxy-CMPF*, tyramine O-sulfate, and unknown X-24337) or anxiety (3-amino-2-piperidone, carotene diol (1), hydroxy-CMPF*, tyramine O-sulfate, unknown X-24337 and X-11315) symptoms after accounting for age, sex, field center, and Hispanic/Latino heritage. Some associations differed by sex. In particular, choline was associated with higher depression and anxiety symptoms in females, but had no association in males. In addition, higher choline remained associated with elevated symptoms of depression after further adjusting for personal SES and health behavioral factors. The resulting MRSs using weights from the discovery dataset were significantly associated with elevated symptoms of depression and anxiety in the replication dataset.

Previous studies have identified lipids and fatty acid metabolism as key metabolic pathways in association with neuropsychiatric health conditions. Our findings further support the existing evidence. For example, hydroxy-CMPF* is linked with lower depression and anxiety symptoms. Hydroxy-CMPF is a hydroxylated metabolite of furan fatty acid metabolites with antioxidant and anti-inflammatory features, and studies have linked it to dietary intake of fatty fish and long-chain polyunsaturated fatty acids.(**39, 40, 41**) The roles of fatty acids (especially omega 3 fatty acids) in cognitive and mental health have been supported by a rich body of evidence in the past decades. Fatty acids can influence various brain processes, including neurotransmission and neuroinflammation, impacting mood and cognitive health.(**42, 43, 44**) In a large Dutch cohort, 34 serum metabolites were associated with risk of major depressive disorder (MDD). Among them, metabolites enriched with long-chain fatty acids were downregulated and metabolites enriched with lysophospholipids were upregulated.(**45**) In another study exploring potential female-specific metabolomic biomarkers for MDD, the researchers identified 18 MDD-related metabolites in the pathways of phospholipids, glycerolipids, and fatty acids, etc.(**46**) In the UK Biobank cohort, Mendelian randomization suggested that fatty acids and intermediate and very large density lipoproteins changed in accordance with disease progression.(**47**) In the Boston Puerto Rican Health Study, acylcarnitines were associated with lower depressive symptoms in 736 Puerto Ricans born in the US 50 states/DC.(**48**) Specifically, acylcarnitines play an important role in the oxidation and transportation of long-chain fatty acids.(**49**) Evidence also showed that there were common alterations in the pathways of arginine and proline metabolism, and alanine, aspartate, and glutamate metabolism in patients with schizophrenia and depression.(**50**) In our study, one of the metabolites we identified in our primary analyses (3-amino-2-piperidone in association with anxiety) is involved in arginine and proline metabolism. It is a derivative of ornithine and is associated with both ornithine and interleukin-8 (IL-8), a small pro-inflammatory cytokine that plays a key role in inflammation and immune responses.(**51**)

Besides the consistent findings with metabolites associated with fatty acids, we identified two new metabolites enriched in vitamin A metabolism (beta-cryptoxanthin and carotene diol (1)) associated with lower psychological distress. Both can be categorized as carotenoids, a group of liposoluble bioactive compounds with strong antioxidant and anti-inflammatory properties.(**52**) Specifically, researchers have recently found in the US National Health and Nutrition Examination Surveys (NHANES), a higher dietary intake of beta cryptoxanthin was associated with less frequent anxiety symptoms in the past month.(**53**) In HCHS/SOL, beta-cryptoxanthin was previously identified to be associated with higher cognitive function.(**54**) It serves as a precursor of vitamin A and is found in tropical and citrus fruits, with an important role in antioxidant defense and cell-to-cell communication.(**55**) Due to conjugated double bonds, it also demonstrates a strong antioxidant capacity to remove different oxygen radicals, and inhibit oxidative damage.(**56**) Carotene diols are unoxygenated carotenoids involved in vitamin A metabolism and were previously observed to have decreased levels in female patients with Alzheimer’s disease.(**57**) However, the relationship between carotenoids and psychological health remains largely unknown and inconclusive. Our study provides evidence on the potential psychological benefits of carotenoids and vitamin A related metabolites.

Tyramine O-sulfate participates in tyrosine metabolism, and is a biochemical compound formed when tyramine undergoes sulfation.(**58**) Although direct evidence for a neuropsychiatric role of tyramine o-sulfate remains limited, studies have confirmed the associations of tyrosine with mental health, including depression, stress and cognitive functioning.(**59, 60, 61, 62**) In a psychiatric primary care clinic based study of 34 MDD patients and 31 mentally healthy controls, plasma tyrosine level was significantly lower among cases versus healthy controls.(**61**) In addition, tyrosine intake was associated with reduced cognitive stress based on a literature review of 10 clinical trials (all showed a positive effect).(**59**) In a prior study looking at plasma metabolites and cognitive functioning, we have also reported that tyramine O-sulfate was associated with reduced cognitive functioning (usually in high correlation with late life depressive symptoms) among Hispanic/Latino adults with Puerto Rican heritage in HCHS/SOL.(**54**) The neuropsychiatric role of tyramine O-sulfate requires further investigation with large longitudinal observational cohort data in a diverse population.

Another interesting finding is the effect modification by sex, particularly for choline. In our study, higher choline was associated with higher depression and anxiety symptoms particularly for females. This indicates a potential sex interaction with the effect of choline on psychological health. However, the harmful psychological effects of choline in females were in contrast with the majority of the existing evidence where researchers observed more of its null or protective effects on neuropsychiatric health outcomes.(**63, 64, 65**) In the Hordaland Health Study including a group of middle aged and older adults, plasma choline concentrations were negatively associated with anxiety symptoms but not with depression symptoms.(**63**) Besides, an L-shaped dose-response relationship between dietary choline and depressive symptoms was reported in 12,906 U.S. adults from NHANES 2011 to 2018.(**65**) In that study, depression symptoms sharply decreased with increased dietary choline intake and the decline plateaued when the intake reached 230 mg/d or more. Possible reasons for this discrepancy might be due to the cohort difference (we focused on Hispanic/Latino adults), modeling approach (linear vs. nonlinear, metabolite transformation approach), covariates used for adjustment and type of choline assessment (serum measurement vs. dietary intake). It should be noted that choline is an essential nutrient required for normal development and functioning of the brain. It acts as a precursor for the synthesis of the neurotransmitter acetylcholine, and also participates in the gene modification and regulation associated with brain functioning.(**66**) The role of choline in psychiatric disorders and mood regulation remains unclear and warrants more investigation.

However, our study provides early evidence suggesting that lower blood choline level is associated with elevated mood symptoms in Hispanic/Latino females, but not males. It should be noted that there are a handful of intersex persons in the cohort whose sex chromosomes or reproductive organs do not match the sex identified in the study. For the analysis, they were included and grouped by their identified sex. With the affected number being small, this does not likely bias our overall sex-stratified findings. Further investigation into the underlying dose-response relationship and whether there is truly a harmful effect threshold in the higher range of choline exposure is needed.

Out study has several strengths. First, it is the first study to identify serum metabolites associated with depression and anxiety symptoms in a multi-center US-based Hispanic/Latino adult population. In addition, we identified effect modification by biological sex with limited evidence from the past literature. The MRSs we established for depression and anxiety symptoms are based on multiple identified metabolites from a metabolome-wide LASSO regression (58 for depression and 70 for anxiety symptoms), and have high correlation with the outcomes. However, this study is also subject to some limitations. First, the analysis is cross-sectional. Potential bias from reverse causation or residual confounding is possible. Second, the study is limited to Hispanic/Latino adults who resided within four metropolitan areas (Chicago, IL; Miami, FL; Bronx, NY; San Diego, CA). Therefore, the findings may have limited generalizability to Hispanic/Latino individuals in other regions and countries. Lastly, due to the limited power of analysis for sex-stratified samples, we ran sex-stratified analyses in combined batches with limited replication across the two batches. Future studies with larger sample size and more separate collection time for metabolites and psychological outcomes in a longitudinal setting that facilitates the establishment of causality are needed.

In summary, we identified several metabolites in relation with symptoms of depression and anxiety in Hispanic/Latino individuals with effect modification by sex. Key metabolic pathways involved included arginine and proline metabolism, vitamin A, phospholipid, fatty acid and tyrosine metabolism. Our study also provides new evidence on the potential psychological benefits of carotenoids and vitamin A related metabolites. Future research studying the role of healthy nutritional intervention (such as via taking food rich in carotenoids and vitamin A) for the management of mental health is needed. The metabolite risk scores we established with depression and anxiety in HCHS/SOL hold the potential for serving as earlier and more objective psychological distress screening tools in addition to traditional risk factors for Hispanic/Latino adults.

## Data Availability

Data will be made available upon approval of a manuscript proposal by the HCHS/SOL publication committee, followed by an executive data use agreement between the institution of the applicant and the HCHS/SOL Data Coordinating Center (DCC). Once the data is integrated into the HCHS/SOL dataset on BioLINCC and dbGaP, investigators would also be able to apply for access via these channels.

## Acknowledgements

This work was supported by National Heart, Lung, and Blood Institute (NHLBI) grant R01HL161012 to TS and National Institute on Aging (NIA) grant R01AG80598 to TS. Support for metabolomics profiling was provided by the Junior League of Houston (JLH) Foundation (Houston, Texas) and NHLBI grant R01HL141824 to BY. The authors thank the staff and participants of HCHS/SOL for their important contributions. Investigators website: http://www.cscc.unc.edu/hchs/. The Hispanic Community Health Study/Study of Latinos is a collaborative study supported by contracts from the National Heart, Lung, and Blood Institute (NHLBI) to the University of North Carolina (HHSN268201300001I/N01-HC-65233), University of Miami (HHSN268201300004I/N01-HC-65234), Albert Einstein College of Medicine (HHSN268201300002I/N01-HC-65235), University of Illinois at Chicago (HHSN268201300003I/N01-HC-65236 Northwestern Univ), and San Diego State University (HHSN268201300005I/N01-HC-65237). The following Institutes/Centers/ Offices have contributed to the HCHS/SOL through a transfer of funds to the NHLBI: National Institute on Minority Health and Health Disparities, National Institute on Deafness and Other Communication Disorders, National Institute of Dental and Craniofacial Research, National Institute of Diabetes and Digestive and Kidney Diseases, National Institute of Neurological Disorders and Stroke, NIH Institution-Office of Dietary Supplements.

