## Supplemental materials for "Serum Metabolites Associated with Depression and Anxiety in the Hispanic Community Health Study/Study of Latinos"

**in the Hispanic Community Health Study/Study of Latinos (HCHS/SOL):**

**Supplementary Materials**

**Table of Contents**

Figure S1. Pearson Correlations Across the Identified 8 Metabolites Associated with Depression and Anxiety Symptoms in HCHS/SOL.

Figure S2. Sex-Stratified Forest Plots of Significant Metabolites (Either for Female or Male) Associated with Depression and Anxiety Symptoms Using Batch Combined Baseline Sample. *(FDR p-value < 0.05)*

Figure S3. Forest Plots of Significant Metabolites Associated with Depression and Anxiety Symptoms in HCHS/SOL from Batch-combined Analyses (Overall). *(FDR p-value < 0.05)*.

Table S1. Metabolites Associated with Depression Symptoms by Sex Using Batch-combined Sample at Baseline. *(FDR p-value < 0.05).*

Table S2. Metabolites Associated with Anxiety Symptoms by Sex Using Batch-combined Sample at Baseline. *(FDR p-value < 0.05).*

Table S3. Metabolites Associated with Depression and Anxiety Symptoms from LASSO regression.

Table S4. Associations between Identified Metabolites Associated with Depression and Anxiety Symptoms from Primary Discovery – Replication Analyses Restricting to Participants Without Antidepressant or Anti-anxiety Medication Use at Baseline *(FDR p-value < 0.05, Batch 1).*

Table S5. Metabolites Associated with Depression Symptoms from Batch Combined Study Sample at Baseline *(FDR p-value < 0.05).*

Table S6. Metabolites Associated with Anxiety Symptoms from Batch Combined Study Sample at Baseline *(FDR p-value < 0.05).*

Table S7. Associations between Identified Metabolites and Depression Symptoms from Longitudinal Mixed Effects Model.

Table S8. Causal Effects of Selected Metabolites on Depression and Anxiety Symptoms from One-Sample Mendelian Randomization.

**Figure S1. Pearson Correlations Across the Identified 8 Metabolites Associated with Depression and Anxiety Symptoms in HCHS/SOL.**

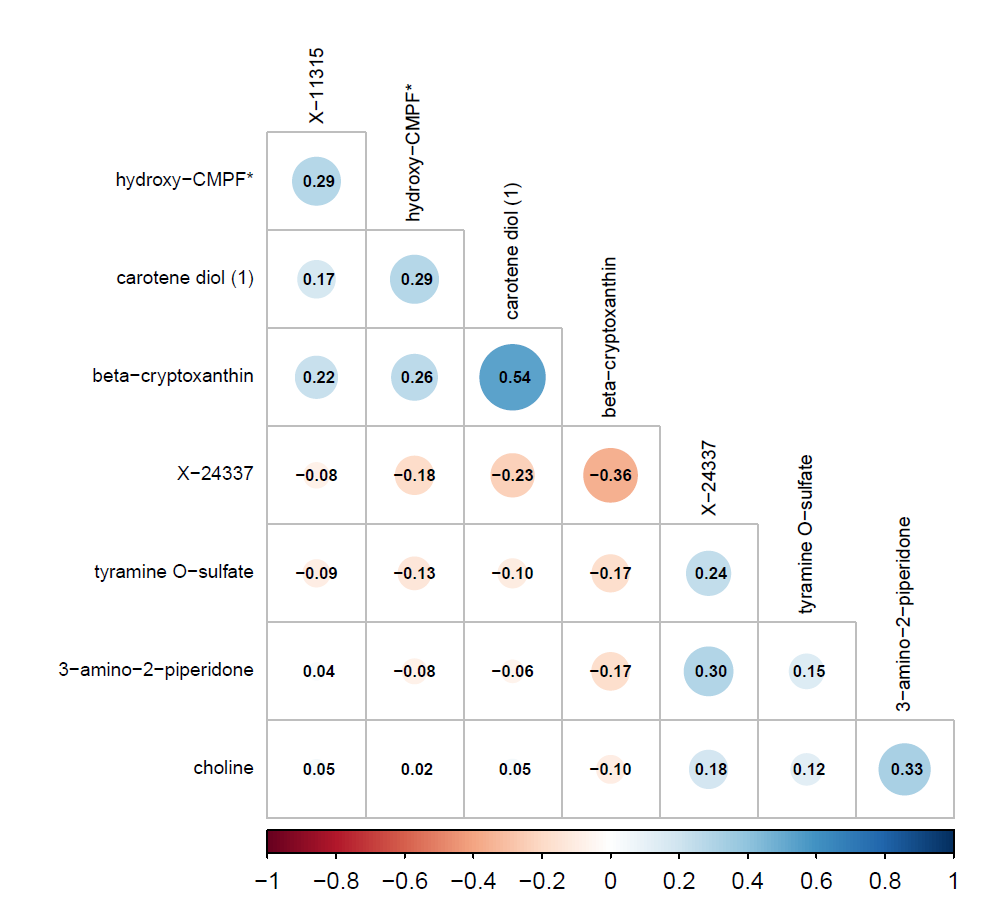

**Figure S2. Sex-Stratified Forest Plots of Significant Metabolites (Either for Female or Male) Associated with Depression and Anxiety Symptoms Using Batch Combined Baseline Sample. *(FDR p-value < 0.05)***

A. **Depression	B. Anxiety**

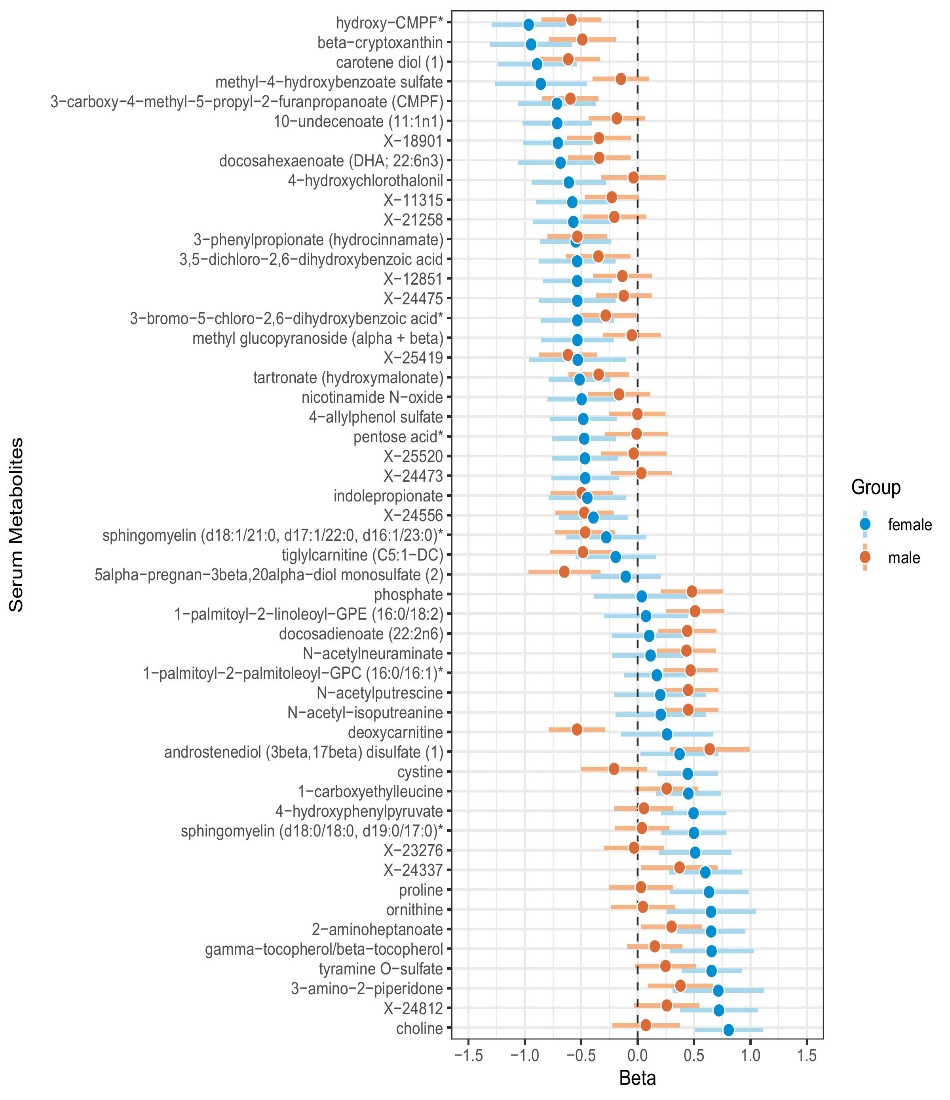

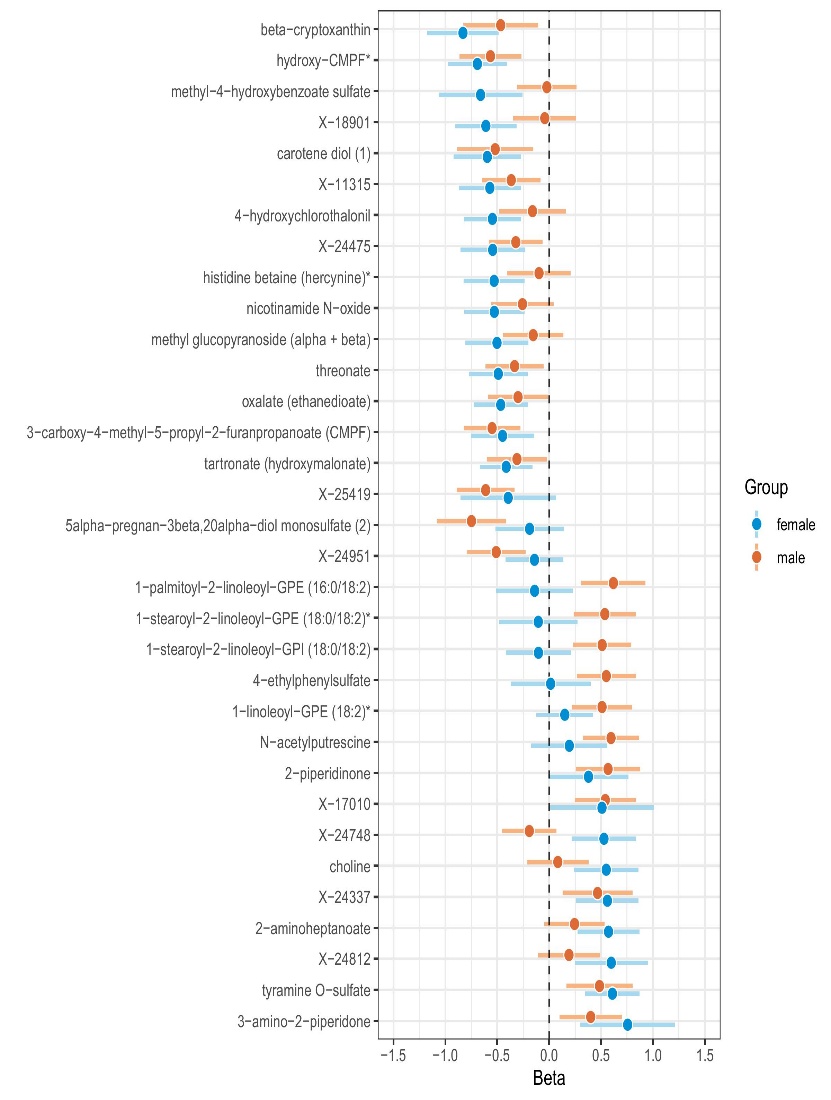

**Figure S3. Forest Plots of Significant Metabolites Associated with Depression and Anxiety Symptoms in HCHS/SOL from Batch-combined Analyses (Overall). *(FDR p-value < 0.05)***

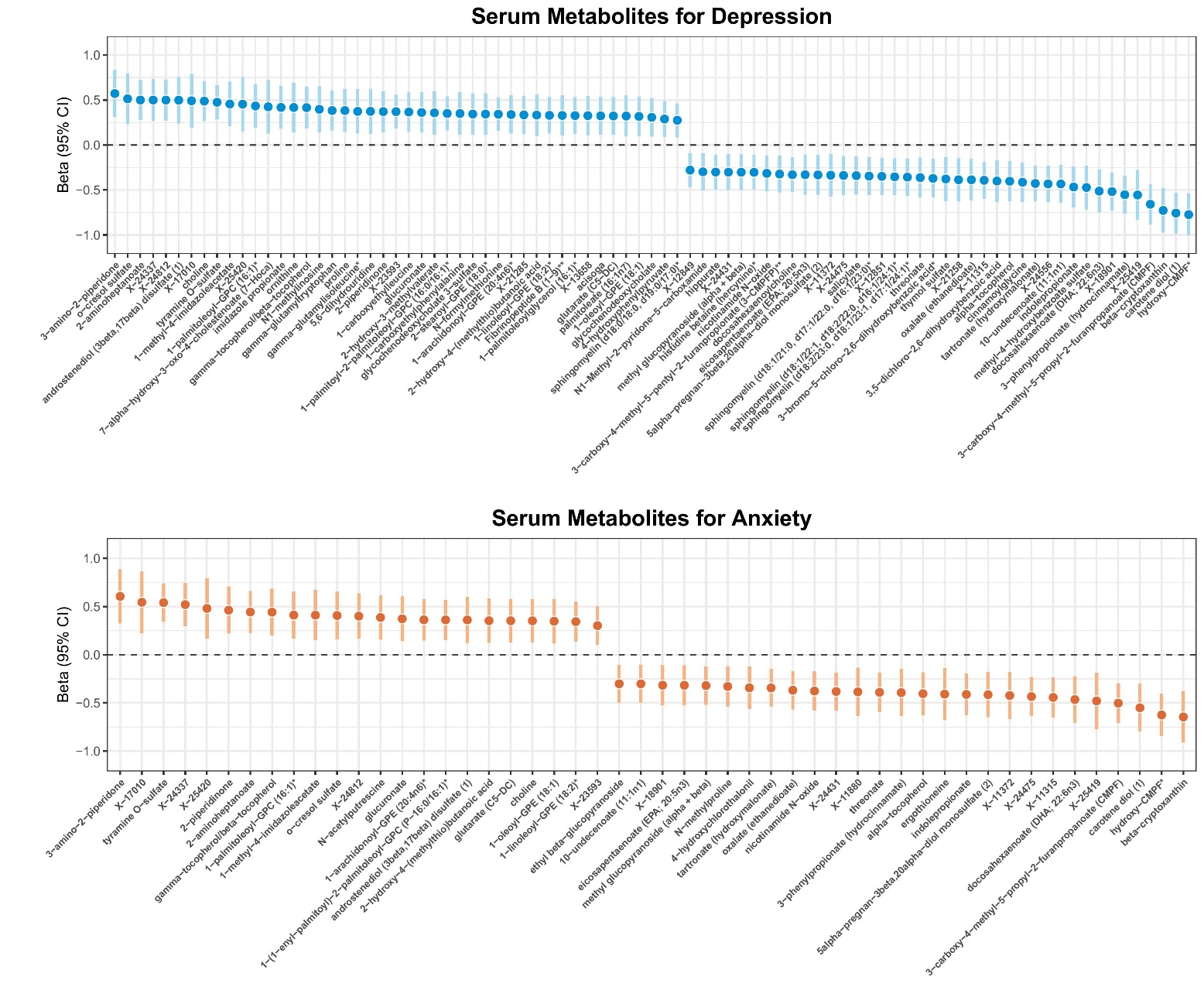

**Table S1. Metabolites Associated with Depression Symptoms by Sex Using Batch-combined Sample at Baseline. *(FDR p-value < 0.05)*** Estimate indicates changes in CES-D-10 score per 1 SD increase in rank-normalized metabolite adjusting for age, sex, field center and Hispanic/Latino background. N/A indicates missing. 95% CI represents 95% confidence intervals.

| **Sex** | **Metabolite** | **Estimate** | **95% CI** | **p** | **FDR p** | **Super Pathway** | **Sub Pathway** |
| --- | --- | --- | --- | --- | --- | --- | --- |
| Females | hydroxy-CMPF* | -0.97 | (-1.29, -0.64) | <0.0001 | <0.0001 | Lipid | Fatty Acid, Dicarboxylate |
|  | choline | 0.81 | (0.51, 1.11) | <0.0001 | 0.0001 | Lipid | Phospholipid Metabolism |
|  | beta-cryptoxanthin | -0.94 | (-1.31, -0.58) | <0.0001 | 0.0001 | Cofactors and Vitamins | Vitamin A Metabolism |
|  | carotene diol (1) | -0.89 | (-1.24, -0.54) | <0.0001 | 0.0001 | Cofactors and Vitamins | Vitamin A Metabolism |
|  | tyramine O-sulfate | 0.66 | (0.39, 0.92) | <0.0001 | 0.0002 | Amino Acid | Tyrosine Metabolism |
|  | 10-undecenoate (11:1n1) | -0.71 | (-1.02, -0.41) | <0.0001 | 0.0007 | Lipid | Medium Chain Fatty Acid |
|  | X-18901 | -0.71 | (-1.01, -0.40) | <0.0001 | 0.0009 | N/A | N/A |
|  | 2-aminoheptanoate | 0.65 | (0.35, 0.95) | <0.0001 | 0.0028 | Lipid | Fatty Acid, Amino |
|  | methyl-4-hydroxybenzoate sulfate | -0.86 | (-1.26, -0.45) | <0.0001 | 0.0031 | Xenobiotics | Benzoate Metabolism |
|  | X-24812 | 0.72 | (0.38, 1.06) | <0.0001 | 0.0031 | N/A | N/A |
|  | 3-carboxy-4-methyl-5-propyl-2-furanpropanoate (CMPF) | -0.71 | (-1.06, -0.37) | 0.0001 | 0.0035 | Lipid | Fatty Acid, Dicarboxylate |
|  | tartronate (hydroxymalonate) | -0.51 | (-0.79, -0.24) | 0.0002 | 0.0152 | Xenobiotics | Food Component/Plant |
|  | X-24337 | 0.60 | (0.28, 0.92) | 0.0003 | 0.0162 | N/A | N/A |
|  | 4-hydroxychlorothalonil | -0.61 | (-0.94, -0.28) | 0.0003 | 0.0173 | Xenobiotics | Chemical |
|  | proline | 0.63 | (0.29, 0.98) | 0.0004 | 0.0188 | Amino Acid | Urea cycle; Arginine and Proline Metabolism |
|  | docosahexaenoate (DHA; 22:6n3) | -0.68 | (-1.06, -0.31) | 0.0004 | 0.0188 | Lipid | Long Chain Polyunsaturated Fatty Acid (n3 and n6) |
|  | X-11315 | -0.58 | (-0.90, -0.26) | 0.0005 | 0.0213 | N/A | N/A |
|  | 3-amino-2-piperidone | 0.72 | (0.31, 1.12) | 0.0006 | 0.023 | Amino Acid | Urea cycle; Arginine and Proline Metabolism |
|  | gamma-tocopherol/beta-tocopherol | 0.66 | (0.28, 1.03) | 0.0006 | 0.023 | Cofactors and Vitamins | Tocopherol Metabolism |
|  | 3-phenylpropionate (hydrocinnamate) | -0.55 | (-0.86, -0.24) | 0.0006 | 0.023 | Xenobiotics | Benzoate Metabolism |
|  | X-12851 | -0.54 | (-0.84, -0.23) | 0.0006 | 0.023 | N/A | N/A |
|  | sphingomyelin (d18:0/18:0, d19:0/17:0)* | 0.50 | (0.21, 0.79) | 0.0007 | 0.023 | Lipid | Dihydrosphingomyelins |
|  | 4-hydroxyphenylpyruvate | 0.50 | (0.21, 0.78) | 0.0008 | 0.027 | Amino Acid | Tyrosine Metabolism |
|  | pentose acid* | -0.47 | (-0.76, -0.19) | 0.0011 | 0.0339 | Partially Characterized Molecules | Partially Characterized Molecules |
|  | cystine | 0.44 | (0.18, 0.71) | 0.0011 | 0.0339 | Amino Acid | Methionine, Cysteine, SAM and Taurine Metabolism |
|  | methyl glucopyranoside (alpha + beta) | -0.53 | (-0.86, -0.21) | 0.0011 | 0.0339 | Xenobiotics | Food Component/Plant |
|  | ornithine | 0.65 | (0.26, 1.04) | 0.0013 | 0.036 | Amino Acid | Urea cycle; Arginine and Proline Metabolism |
|  | 3-bromo-5-chloro-2,6-dihydroxybenzoic acid* | -0.53 | (-0.86, -0.21) | 0.0013 | 0.036 | Xenobiotics | Chemical |
|  | 4-allylphenol sulfate | -0.48 | (-0.78, -0.19) | 0.0014 | 0.0365 | Xenobiotics | Food Component/Plant |
|  | nicotinamide N-oxide | -0.50 | (-0.80, -0.19) | 0.0014 | 0.0365 | Cofactors and Vitamins | Nicotinate and Nicotinamide Metabolism |
|  | X-25520 | -0.47 | (-0.76, -0.18) | 0.0017 | 0.0413 | N/A | N/A |
|  | X-23276 | 0.51 | (0.19, 0.83) | 0.0018 | 0.0436 | N/A | N/A |
|  | X-21258 | -0.57 | (-0.93, -0.21) | 0.0019 | 0.0436 | N/A | N/A |
|  | 1-carboxyethylleucine | 0.45 | (0.17, 0.73) | 0.002 | 0.0436 | N/A | N/A |
|  | X-24475 | -0.54 | (-0.87, -0.20) | 0.002 | 0.0436 | N/A | N/A |
|  | 3,5-dichloro-2,6-dihydroxybenzoic acid | -0.54 | (-0.88, -0.20) | 0.002 | 0.0436 | Xenobiotics | Chemical |
|  | X-24473 | -0.47 | (-0.76, -0.17) | 0.0024 | 0.0492 | N/A | N/A |
| Males | X-25419 | -0.62 | (-0.87, -0.36) | <0.0001 | 0.0012 | N/A | N/A |
|  | 3-carboxy-4-methyl-5-propyl-2-furanpropanoate (CMPF) | -0.60 | (-0.84, -0.35) | <0.0001 | 0.0012 | Lipid | Fatty Acid, Dicarboxylate |
|  | hydroxy-CMPF* | -0.59 | (-0.85, -0.32) | <0.0001 | 0.0035 | Lipid | Fatty Acid, Dicarboxylate |
|  | carotene diol (1) | -0.62 | (-0.89, -0.34) | <0.0001 | 0.0035 | Cofactors and Vitamins | Vitamin A Metabolism |
|  | deoxycarnitine | -0.54 | (-0.79, -0.29) | <0.0001 | 0.0044 | Lipid | Carnitine Metabolism |
|  | 5alpha-pregnan-3beta,20alpha-diol monosulfate (2) | -0.65 | (-0.97, -0.33) | 0.0001 | 0.009 | Lipid | Progestin Steroids |
|  | 3-phenylpropionate (hydrocinnamate) | -0.53 | (-0.80, -0.27) | 0.0001 | 0.0092 | Xenobiotics | Benzoate Metabolism |
|  | 1-palmitoyl-2-linoleoyl-GPE (16:0/18:2) | 0.51 | (0.25, 0.76) | 0.0001 | 0.0101 | Lipid | Phosphatidylethanolamine (PE) |
|  | 1-palmitoyl-2-palmitoleoyl-GPC (16:0/16:1)* | 0.47 | (0.23, 0.71) | 0.0001 | 0.0123 | Lipid | Phosphatidylcholine (PC) |
|  | X-24556 | -0.47 | (-0.73, -0.21) | 0.0004 | 0.0279 | N/A | N/A |
|  | indolepropionate | -0.50 | (-0.77, -0.22) | 0.0004 | 0.0279 | Amino Acid | Tryptophan Metabolism |
|  | androstenediol (3beta,17beta) disulfate (1) | 0.64 | (0.29, 0.99) | 0.0004 | 0.0279 | Lipid | Androgenic Steroids |
|  | sphingomyelin (d18:1/21:0, d17:1/22:0, d16:1/23:0)* | -0.47 | (-0.73, -0.20) | 0.0006 | 0.0359 | Lipid | Sphingomyelins |
|  | phosphate | 0.48 | (0.21, 0.76) | 0.0007 | 0.0359 | Energy | Oxidative Phosphorylation |
|  | docosadienoate (22:2n6) | 0.44 | (0.18, 0.70) | 0.0008 | 0.0429 | Lipid | Long Chain Polyunsaturated Fatty Acid (n3 and n6) |
|  | N-acetyl-isoputreanine | 0.45 | (0.19, 0.71) | 0.0009 | 0.0429 | Amino Acid | Polyamine Metabolism |
|  | N-acetylputrescine | 0.45 | (0.18, 0.71) | 0.001 | 0.0459 | Amino Acid | Polyamine Metabolism |
|  | beta-cryptoxanthin | -0.49 | (-0.78, -0.20) | 0.0012 | 0.0499 | Cofactors and Vitamins | Vitamin A Metabolism |
|  | tiglylcarnitine (C5:1-DC) | -0.49 | (-0.78, -0.19) | 0.0012 | 0.0499 | Amino Acid | Leucine, Isoleucine and Valine Metabolism |
|  | N-acetylneuraminate | 0.43 | (0.17, 0.70) | 0.0013 | 0.0499 | Carbohydrate | Amino sugar Metabolism |

**Table S2. Metabolites Associated with Anxiety Symptoms by Sex Using Batch-combined Sample at Baseline. *(FDR p-value < 0.05)*** Estimate indicates changes in STAI-10 score per 1 SD increase in rank-normalized metabolite adjusting for age, sex, field center and Hispanic/Latino background. N/A indicates missing. 95% CI represents 95% confidence intervals.

| **Sex** | **Metabolite** | **Estimate** | **95% CI** | **p** | **FDR p** | **Super Pathway** | **Sub Pathway** |
| --- | --- | --- | --- | --- | --- | --- | --- |
| Females | hydroxy-CMPF* | -0.69 | (-0.97, -0.41) | <0.0001 | 0.0009 | Lipid | Fatty Acid, Dicarboxylate |
|  | beta-cryptoxanthin | -0.83 | (-1.17, -0.49) | <0.0001 | 0.0009 | Cofactors and Vitamins | Vitamin A Metabolism |
|  | tyramine O-sulfate | 0.61 | (0.35, 0.87) | <0.0001 | 0.0015 | Amino Acid | Tyrosine Metabolism |
|  | X-18901 | -0.61 | (-0.90, -0.32) | 0.0001 | 0.0098 | N/A | N/A |
|  | 4-hydroxychlorothalonil | -0.55 | (-0.82, -0.27) | 0.0001 | 0.0153 | Xenobiotics | Chemical |
|  | X-11315 | -0.57 | (-0.87, -0.28) | 0.0002 | 0.0211 | N/A | N/A |
|  | 2-aminoheptanoate | 0.57 | (0.27, 0.87) | 0.0002 | 0.0211 | Lipid | Fatty Acid, Amino |
|  | X-24337 | 0.56 | (0.26, 0.86) | 0.0003 | 0.0249 | N/A | N/A |
|  | carotene diol (1) | -0.60 | (-0.92, -0.27) | 0.0003 | 0.0281 | Cofactors and Vitamins | Vitamin A Metabolism |
|  | nicotinamide N-oxide | -0.53 | (-0.82, -0.24) | 0.0004 | 0.0283 | Cofactors and Vitamins | Nicotinate and Nicotinamide Metabolism |
|  | histidine betaine (hercynine)* | -0.53 | (-0.82, -0.24) | 0.0004 | 0.0294 | Xenobiotics | Food Component/Plant |
|  | oxalate (ethanedioate) | -0.47 | (-0.73, -0.21) | 0.0005 | 0.0295 | Cofactors and Vitamins | Ascorbate and Aldarate Metabolism |
|  | choline | 0.55 | (0.24, 0.86) | 0.0005 | 0.0321 | Lipid | Phospholipid Metabolism |
|  | X-24475 | -0.54 | (-0.85, -0.23) | 0.0006 | 0.0339 | N/A | N/A |
|  | threonate | -0.49 | (-0.77, -0.21) | 0.0007 | 0.0361 | Cofactors and Vitamins | Ascorbate and Aldarate Metabolism |
|  | X-24748 | 0.53 | (0.22, 0.84) | 0.0008 | 0.0395 | N/A | N/A |
|  | X-24812 | 0.60 | (0.25, 0.95) | 0.0009 | 0.0395 | N/A | N/A |
|  | methyl glucopyranoside (alpha + beta) | -0.50 | (-0.80, -0.20) | 0.0012 | 0.0475 | Xenobiotics | Food Component/Plant |
|  | 3-amino-2-piperidone | 0.76 | (0.30, 1.21) | 0.0012 | 0.0475 | Amino Acid | Urea cycle; Arginine and Proline Metabolism |
|  | methyl-4-hydroxybenzoate sulfate | -0.66 | (-1.06, -0.26) | 0.0013 | 0.0485 | Xenobiotics | Benzoate Metabolism |
|  | tartronate (hydroxymalonate) | -0.41 | (-0.67, -0.16) | 0.0013 | 0.0485 | Xenobiotics | Food Component/Plant |
| Males | 5alpha-pregnan-3beta,20alpha-diol monosulfate (2) | -0.75 | (-1.08, -0.42) | <0.0001 | 0.0047 | Lipid | Progestin Steroids |
|  | N-acetylputrescine | 0.60 | (0.33, 0.86) | <0.0001 | 0.0047 | Amino Acid | Polyamine Metabolism |
|  | X-25419 | -0.61 | (-0.89, -0.33) | <0.0001 | 0.0047 | N/A | N/A |
|  | 3-carboxy-4-methyl-5-propyl-2-furanpropanoate (CMPF) | -0.55 | (-0.82, -0.28) | 0.0001 | 0.0141 | Lipid | Fatty Acid, Dicarboxylate |
|  | 1-palmitoyl-2-linoleoyl-GPE (16:0/18:2) | 0.62 | (0.31, 0.93) | 0.0001 | 0.0141 | Lipid | Phosphatidylethanolamine (PE) |
|  | 4-ethylphenylsulfate | 0.55 | (0.27, 0.83) | 0.0001 | 0.0187 | Xenobiotics | Benzoate Metabolism |
|  | hydroxy-CMPF* | -0.57 | (-0.86, -0.27) | 0.0002 | 0.0214 | Lipid | Fatty Acid, Dicarboxylate |
|  | 2-piperidinone | 0.57 | (0.26, 0.87) | 0.0003 | 0.0291 | Xenobiotics | Food Component/Plant |
|  | X-17010 | 0.54 | (0.25, 0.83) | 0.0003 | 0.0291 | N/A | N/A |
|  | 1-stearoyl-2-linoleoyl-GPI (18:0/18:2) | 0.51 | (0.23, 0.79) | 0.0004 | 0.0291 | Lipid | Phosphatidylinositol (PI) |
|  | X-24951 | -0.51 | (-0.79, -0.23) | 0.0004 | 0.0291 | N/A | N/A |
|  | 1-stearoyl-2-linoleoyl-GPE (18:0/18:2)* | 0.54 | (0.24, 0.83) | 0.0005 | 0.0291 | Lipid | Phosphatidylethanolamine (PE) |
|  | 1-linoleoyl-GPE (18:2)* | 0.51 | (0.22, 0.80) | 0.0005 | 0.0322 | Lipid | Lysophospholipid |

**Table S3. Metabolites Associated with Depression and Anxiety Symptoms from LASSO regression.** Shaded entries (34 metabolites) indicate overlapping metabolites associated with depression and anxiety symptoms. N/A indicates missing.

| **Order** | **CESD10 Metabolite** | **CESD10 Coefficient** | **STAI10 Metabolite** | **STAI10 Coefficient** |
| --- | --- | --- | --- | --- |
| 1 | 3-phenylpropionate (hydrocinnamate) | -0.23 | N/A | N/A |
| 2 | hippurate | -0.03 | hippurate | -0.18 |
| 3 | glucuronate | 0.06 | glucuronate | 0.11 |
| 4 | N/A | N/A | glycerol 3-phosphate | -0.05 |
| 5 | theobromine | -0.01 | theobromine | -0.05 |
| 6 | sphingosine 1-phosphate | 0.02 | N/A | N/A |
| 7 | 1,2-dipalmitoyl-GPC (16:0/16:0) | 0.13 | 1,2-dipalmitoyl-GPC (16:0/16:0) | 0.15 |
| 8 | docosahexaenoate (DHA; 22:6n3) | -0.05 | N/A | N/A |
| 9 | indolepropionate | -0.04 | indolepropionate | -0.14 |
| 10 | 3-carboxy-4-methyl-5-propyl-2-furanpropanoate (CMPF) | -0.12 | N/A | N/A |
| 11 | 10-undecenoate (11:1n1) | -0.19 | 10-undecenoate (11:1n1) | -0.23 |
| 12 | 1-methyl-4-imidazoleacetate | 0.05 | 1-methyl-4-imidazoleacetate | 0.01 |
| 13 | piperine | 0.00 | piperine | -0.04 |
| 14 | N/A | N/A | gamma-glutamylglycine | 0.15 |
| 15 | N/A | N/A | gamma-glutamyltryptophan | 0.02 |
| 16 | phenylacetylglutamine | 0.10 | phenylacetylglutamine | 0.04 |
| 17 | 5-methyluridine (ribothymidine) | -0.10 | 5-methyluridine (ribothymidine) | -0.04 |
| 18 | 1-palmitoleoyl-GPC (16:1)* | 0.09 | N/A | N/A |
| 19 | 2-hydroxypalmitate | 0.00 | N/A | N/A |
| 20 | N/A | N/A | isobutyrylglycine | -0.01 |
| 21 | 7-alpha-hydroxy-3-oxo-4-cholestenoate (7-Hoca) | 0.13 | 7-alpha-hydroxy-3-oxo-4-cholestenoate (7-Hoca) | 0.26 |
| 22 | N/A | N/A | 3,4-dihydroxybutyrate | 0.06 |
| 23 | tryptophan betaine | -0.09 | tryptophan betaine | -0.05 |
| 24 | 1-palmitoyl-2-linoleoyl-GPE (16:0/18:2) | 0.01 | N/A | N/A |
| 25 | N/A | N/A | 5alpha-pregnan-3beta,20alpha-diol monosulfate (2) | -0.03 |
| 26 | 5alpha-androstan-3beta,17alpha-diol disulfate | -0.03 | 5alpha-androstan-3beta,17alpha-diol disulfate | -0.04 |
| 27 | andro steroid monosulfate C19H28O6S (1)* | 0.01 | N/A | N/A |
| 28 | nicotinamide N-oxide | -0.05 | nicotinamide N-oxide | -0.13 |
| 29 | eicosanedioate (C20-DC) | -0.02 | eicosanedioate (C20-DC) | -0.10 |
| 30 | N/A | N/A | leucylglycine | 0.18 |
| 31 | N/A | N/A | trimethylamine N-oxide | 0.07 |
| 32 | N/A | N/A | alliin | -0.03 |
| 33 | 2-stearoyl-GPE (18:0)* | 0.04 | 2-stearoyl-GPE (18:0)* | 0.01 |
| 34 | N/A | N/A | N-acetylcarnosine | -0.23 |
| 35 | N/A | N/A | histidine betaine (hercynine)* | 0.00 |
| 36 | 3-methyl catechol sulfate (1) | 0.01 | N/A | N/A |
| 37 | N/A | N/A | 2-aminoheptanoate | 0.12 |
| 38 | N/A | N/A | N2,N5-diacetylornithine | -0.16 |
| 39 | O-sulfo-L-tyrosine | -0.01 | O-sulfo-L-tyrosine | -0.04 |
| 40 | N/A | N/A | 2-keto-3-deoxy-gluconate | -0.03 |
| 41 | tyramine O-sulfate | 0.07 | tyramine O-sulfate | 0.10 |
| 42 | glycodeoxycholate 3-sulfate | 0.04 | N/A | N/A |
| 43 | N/A | N/A | 1-stearoyl-2-arachidonoyl-GPE (18:0/20:4) | 0.11 |
| 44 | sphingomyelin (d18:0/18:0, d19:0/17:0)* | 0.05 | sphingomyelin (d18:0/18:0, d19:0/17:0)* | 0.04 |
| 45 | 1-(1-enyl-palmitoyl)-2-palmitoleoyl-GPC (P-16:0/16:1)* | 0.02 | 1-(1-enyl-palmitoyl)-2-palmitoleoyl-GPC (P-16:0/16:1)* | 0.09 |
| 46 | 2'-O-methyluridine | 0.10 | 2'-O-methyluridine | 0.09 |
| 47 | sphingomyelin (d18:2/18:1)* | 0.02 | N/A | N/A |
| 48 | N/A | N/A | sphingomyelin (d18:2/24:2)* | 0.01 |
| 49 | carotene diol (1) | -0.27 | carotene diol (1) | -0.10 |
| 50 | N/A | N/A | hydroxy-CMPF* | -0.03 |
| 51 | N/A | N/A | octadecadienedioate (C18:2-DC)* | 0.02 |
| 52 | 3-carboxy-4-methyl-5-pentyl-2-furanpropionate (3-CMPFP)** | -0.15 | 3-carboxy-4-methyl-5-pentyl-2-furanpropionate (3-CMPFP)** | -0.07 |
| 53 | 3-amino-2-piperidone | 0.11 | 3-amino-2-piperidone | 0.14 |
| 54 | N/A | N/A | ascorbic acid 3-sulfate* | 0.04 |
| 55 | N/A | N/A | N2-acetyl,N6,N6-dimethyllysine | -0.08 |
| 56 | N/A | N/A | lithocholate sulfate (1) | -0.06 |
| 57 | hydroxypalmitoyl sphingomyelin (d18:1/16:0(OH))** | 0.04 | hydroxypalmitoyl sphingomyelin (d18:1/16:0(OH))** | 0.06 |
| 58 | 3,5-dichloro-2,6-dihydroxybenzoic acid | -0.13 | N/A | N/A |
| 59 | 1-methyl-5-imidazolelactate | -0.13 | 1-methyl-5-imidazolelactate | -0.06 |
| 60 | N/A | N/A | 3-bromo-5-chloro-2,6-dihydroxybenzoic acid* | -0.01 |
| 61 | 3-hydroxyoctanoylcarnitine (2) | -0.01 | N/A | N/A |
| 62 | N/A | N/A | bilirubin degradation product, C17H20N2O5 (1)** | -0.04 |
| 63 | N/A | N/A | N-acetylvaline | -0.09 |
| 64 | N/A | N/A | alpha-tocopherol | -0.14 |
| 65 | 3-hydroxyisobutyrate | -0.11 | N/A | N/A |
| 66 | N/A | N/A | 4-acetamidobutanoate | 0.04 |
| 67 | N/A | N/A | N-palmitoyl-sphingosine (d18:1/16:0) | 0.00 |
| 68 | N/A | N/A | 9,10-DiHOME | 0.07 |
| 69 | N6,N6,N6-trimethyllysine | -0.05 | N/A | N/A |
| 70 | N/A | N/A | guanidinoacetate | 0.04 |
| 71 | cortisol | -0.01 | N/A | N/A |
| 72 | ornithine | 0.10 | N/A | N/A |
| 73 | N/A | N/A | orotate | -0.02 |
| 74 | phosphate | 0.13 | phosphate | 0.14 |
| 75 | salicylate | -0.01 | N/A | N/A |
| 76 | glutamate | 0.01 | glutamate | 0.00 |
| 77 | N/A | N/A | cysteine | -0.01 |
| 78 | N/A | N/A | arachidate (20:0) | -0.01 |
| 79 | N/A | N/A | X-11880 | -0.03 |
| 80 | X-12117 | 0.03 | N/A | N/A |
| 81 | X-12798 | -0.14 | X-12798 | -0.21 |
| 82 | N/A | N/A | X-12849 | -0.01 |
| 83 | X-13658 | 0.15 | N/A | N/A |
| 84 | X-17010 | 0.19 | X-17010 | 0.24 |
| 85 | X-18901 | -0.09 | N/A | N/A |
| 86 | X-21258 | -0.06 | N/A | N/A |
| 87 | X-21285 | 0.02 | X-21285 | 0.06 |
| 88 | N/A | N/A | X-21286 | -0.06 |
| 89 | X-23276 | 0.00 | N/A | N/A |
| 90 | X-24473 | -0.02 | X-24473 | -0.05 |
| 91 | N/A | N/A | X-24475 | -0.12 |
| 92 | X-24812 | 0.21 | N/A | N/A |
| 93 | X-25419 | -0.04 | X-25419 | -0.08 |
| 94 | X-25420 | 0.16 | X-25420 | 0.20 |

**Table S4. Associations between Identified Metabolites Associated with Depression and Anxiety Symptoms from Primary Discovery – Replication Analyses Restricting to Participants Without Antidepressant or Anti-anxiety Medication Use at Baseline *(FDR p-value < 0.05, Batch 1).*** Results are only shown for significant associations. Estimate indicates changes in CES-D-10 (depression) or STAI-10 (anxiety) continuous score per 1 SD increase in rank-normalized metabolite adjusting for age, sex, field center and Hispanic/Latino background. 95% CI represents 95% confidence intervals.

| **Metabolite** | **Estimate** | **95% CI** | **p** | **FDR p** |
| --- | --- | --- | --- | --- |
| Depression | | | | |
| beta-cryptoxanthin | -0.41 | (-0.68, -0.14) | 0.0031 | 0.0051 |
| tyramine O-sulfate | 0.25 | (0.02, 0.49) | 0.0333 | 0.0417 |
| hydroxy-CMPF* | -0.67 | (-0.93, -0.41) | <0.0001 | <0.0001 |
| choline | 0.34 | (0.12, 0.56) | 0.0025 | 0.0051 |
| Anxiety | | | | |
| tyramine O-sulfate | 0.35 | (0.09, 0.61) | 0.0074 | 0.0149 |
| carotene diol (1) | -0.34 | (-0.63, -0.06) | 0.0186 | 0.0223 |
| hydroxy-CMPF* | -0.57 | (-0.84, -0.31) | <0.0001 | 0.0002 |
| X-11315 | -0.45 | (-0.69, -0.21) | 0.0002 | 0.0007 |
| X-24337 | 0.31 | (0.05, 0.56) | 0.0183 | 0.0223 |

**Table S5. Metabolites Associated with Depression Symptoms from Batch Combined Study Sample at Baseline *(FDR p-value < 0.05).*** Highlighted shaded metabolites indicate overlap with main findings (discovery-replication approach). Estimate indicates changes in CES-D-10 (depression) continuous score per 1 SD increase in rank-normalized metabolite adjusting for age, sex, field center and Hispanic/Latino background. 95% CI represents 95% confidence intervals.

| **Metabolite** | **Estimate** | **95% CI** | **p** | **FDR p** | **Super Pathway** | **Sub Pathway** |
| --- | --- | --- | --- | --- | --- | --- |
| hydroxy-CMPF* | -0.77 | (-1.01, -0.54) | <0.0001 | <0.0001 | Lipid | Fatty Acid, Dicarboxylate |
| carotene diol (1) | -0.76 | (-0.98, -0.53) | <0.0001 | <0.0001 | Cofactors and Vitamins | Vitamin A Metabolism |
| 3-carboxy-4-methyl-5-propyl-2-furanpropanoate (CMPF) | -0.66 | (-0.88, -0.44) | <0.0001 | <0.0001 | Lipid | Fatty Acid, Dicarboxylate |
| beta-cryptoxanthin | -0.73 | (-0.97, -0.48) | <0.0001 | <0.0001 | Cofactors and Vitamins | Vitamin A Metabolism |
| 3-phenylpropionate (hydrocinnamate) | -0.55 | (-0.76, -0.34) | <0.0001 | <0.0001 | Xenobiotics | Benzoate Metabolism |
| X-18901 | -0.52 | (-0.73, -0.31) | <0.0001 | 0.0001 | N/A | N/A |
| tyramine O-sulfate | 0.48 | (0.28, 0.67) | <0.0001 | 0.0001 | Amino Acid | Tyrosine Metabolism |
| 2-aminoheptanoate | 0.50 | (0.28, 0.72) | <0.0001 | 0.0009 | Lipid | Fatty Acid, Amino |
| X-24812 | 0.50 | (0.28, 0.72) | <0.0001 | 0.0012 | N/A | N/A |
| choline | 0.49 | (0.27, 0.71) | <0.0001 | 0.0012 | Lipid | Phospholipid Metabolism |
| 3-amino-2-piperidone | 0.57 | (0.31, 0.83) | <0.0001 | 0.0013 | Amino Acid | Urea cycle; Arginine and Proline Metabolism |
| docosahexaenoate (DHA; 22:6n3) | -0.51 | (-0.75, -0.28) | <0.0001 | 0.0014 | Lipid | Long Chain Polyunsaturated Fatty Acid (n3 and n6) |
| tartronate (hydroxymalonate) | -0.43 | (-0.63, -0.23) | <0.0001 | 0.0014 | Xenobiotics | Food Component/Plant |
| X-24337 | 0.50 | (0.27, 0.73) | <0.0001 | 0.0014 | N/A | N/A |
| X-24556 | -0.43 | (-0.63, -0.23) | <0.0001 | 0.0014 | N/A | N/A |
| indolepropionate | -0.47 | (-0.69, -0.24) | <0.0001 | 0.0022 | Amino Acid | Tryptophan Metabolism |
| 10-undecenoate (11:1n1) | -0.43 | (-0.64, -0.23) | <0.0001 | 0.0022 | Lipid | Medium Chain Fatty Acid |
| X-25419 | -0.56 | (-0.83, -0.28) | 0.0001 | 0.0037 | N/A | N/A |
| X-23593 | 0.37 | (0.19, 0.56) | 0.0001 | 0.0037 | N/A | N/A |
| methyl-4-hydroxybenzoate sulfate | -0.47 | (-0.72, -0.23) | 0.0001 | 0.0055 | Xenobiotics | Benzoate Metabolism |
| androstenediol (3beta,17beta) disulfate (1) | 0.50 | (0.24, 0.76) | 0.0002 | 0.0055 | Lipid | Androgenic Steroids |
| cinnamoylglycine | -0.41 | (-0.63, -0.20) | 0.0002 | 0.0068 | Xenobiotics | Food Component/Plant |
| X-11315 | -0.39 | (-0.60, -0.19) | 0.0002 | 0.0068 | N/A | N/A |
| 1-palmitoyl-2-palmitoleoyl-GPC (16:0/16:1)* | 0.35 | (0.16, 0.54) | 0.0003 | 0.009 | Lipid | Phosphatidylcholine (PC) |
| N-formylmethionine | 0.34 | (0.16, 0.53) | 0.0003 | 0.0092 | Amino Acid | Methionine, Cysteine, SAM and Taurine Metabolism |
| 1-methyl-4-imidazoleacetate | 0.46 | (0.21, 0.70) | 0.0003 | 0.0092 | Amino Acid | Histidine Metabolism |
| o-cresol sulfate | 0.51 | (0.23, 0.79) | 0.0003 | 0.0096 | Xenobiotics | Benzoate Metabolism |
| 1-palmitoleoyl-GPC (16:1)* | 0.43 | (0.20, 0.67) | 0.0004 | 0.0109 | Lipid | Lysophospholipid |
| gamma-tocopherol/beta-tocopherol | 0.42 | (0.18, 0.65) | 0.0005 | 0.0118 | Cofactors and Vitamins | Tocopherol Metabolism |
| alpha-tocopherol | -0.40 | (-0.63, -0.18) | 0.0005 | 0.0118 | Cofactors and Vitamins | Tocopherol Metabolism |
| imidazole propionate | 0.42 | (0.18, 0.66) | 0.0006 | 0.0147 | Amino Acid | Histidine Metabolism |
| gamma-glutamyltryptophan | 0.38 | (0.16, 0.60) | 0.0006 | 0.015 | Peptide | Gamma-glutamyl Amino Acid |
| 3-bromo-5-chloro-2,6-dihydroxybenzoic acid* | -0.37 | (-0.58, -0.16) | 0.0007 | 0.015 | Xenobiotics | Chemical |
| sphingomyelin (d18:2/23:0, d18:1/23:1, d17:1/24:1)* | -0.36 | (-0.56, -0.15) | 0.0007 | 0.015 | Lipid | Sphingomyelins |
| sphingomyelin (d18:1/22:1, d18:2/22:0, d16:1/24:1)* | -0.35 | (-0.56, -0.15) | 0.0007 | 0.015 | Lipid | Sphingomyelins |
| 3,5-dichloro-2,6-dihydroxybenzoic acid | -0.40 | (-0.63, -0.17) | 0.0008 | 0.016 | Xenobiotics | Chemical |
| docosahexaenoylcholine | -0.33 | (-0.52, -0.14) | 0.0008 | 0.016 | Lipid | Fatty Acid Metabolism (Acyl Choline) |
| 1-carboxyethylleucine | 0.37 | (0.15, 0.58) | 0.0008 | 0.0169 | N/A | N/A |
| X-12851 | -0.35 | (-0.55, -0.14) | 0.0009 | 0.0187 | N/A | N/A |
| oxalate (ethanedioate) | -0.39 | (-0.62, -0.16) | 0.001 | 0.019 | Cofactors and Vitamins | Ascorbate and Aldarate Metabolism |
| X-17010 | 0.49 | (0.19, 0.79) | 0.0013 | 0.0233 | N/A | N/A |
| glucuronate | 0.36 | (0.14, 0.58) | 0.0013 | 0.0233 | Carbohydrate | Amino sugar Metabolism |
| sphingomyelin (d18:1/21:0, d17:1/22:0, d16:1/23:0)* | -0.35 | (-0.56, -0.14) | 0.0013 | 0.0233 | Lipid | Sphingomyelins |
| threonate | -0.36 | (-0.58, -0.14) | 0.0014 | 0.0233 | Cofactors and Vitamins | Ascorbate and Aldarate Metabolism |
| 1-linoleoyl-GPE (18:2)* | 0.33 | (0.13, 0.53) | 0.0014 | 0.0233 | Lipid | Lysophospholipid |
| 2-piperidinone | 0.37 | (0.14, 0.60) | 0.0014 | 0.0233 | Xenobiotics | Food Component/Plant |
| salicylate | -0.34 | (-0.55, -0.13) | 0.0015 | 0.0241 | Xenobiotics | Drug - Topical Agents |
| X-21258 | -0.39 | (-0.62, -0.15) | 0.0015 | 0.0241 | N/A | N/A |
| proline | 0.38 | (0.15, 0.62) | 0.0015 | 0.0241 | Amino Acid | Urea cycle; Arginine and Proline Metabolism |
| 1-arachidonoyl-GPE (20:4n6)* | 0.34 | (0.13, 0.55) | 0.0017 | 0.0254 | Lipid | Lysophospholipid |
| 1-palmitoleoylglycerol (16:1)* | 0.33 | (0.12, 0.53) | 0.0017 | 0.026 | Lipid | Monoacylglycerol |
| 3-carboxy-4-methyl-5-pentyl-2-furanpropionate (3-CMPFP)** | -0.32 | (-0.53, -0.12) | 0.0019 | 0.0277 | Lipid | Fatty Acid, Dicarboxylate |
| nicotinamide N-oxide | -0.31 | (-0.51, -0.12) | 0.002 | 0.0288 | Cofactors and Vitamins | Nicotinate and Nicotinamide Metabolism |
| N1-methylinosine | 0.40 | (0.15, 0.65) | 0.002 | 0.0288 | Nucleotide | Purine Metabolism, (Hypo)Xanthine/Inosine |
| acisoga | 0.32 | (0.12, 0.53) | 0.0021 | 0.0288 | Amino Acid | Polyamine Metabolism |
| X-21285 | 0.34 | (0.12, 0.55) | 0.0021 | 0.0288 | N/A | N/A |
| histidine betaine (hercynine)* | -0.30 | (-0.50, -0.11) | 0.0022 | 0.0296 | Xenobiotics | Food Component/Plant |
| X-24475 | -0.34 | (-0.55, -0.12) | 0.0022 | 0.0296 | N/A | N/A |
| glutarate (C5-DC) | 0.32 | (0.12, 0.53) | 0.0024 | 0.0307 | Lipid | Fatty Acid, Dicarboxylate |
| hippurate | -0.30 | (-0.49, -0.11) | 0.0024 | 0.0307 | Xenobiotics | Benzoate Metabolism |
| thymol sulfate | -0.38 | (-0.62, -0.13) | 0.0024 | 0.0307 | Xenobiotics | Food Component/Plant |
| methyl glucopyranoside (alpha + beta) | -0.30 | (-0.50, -0.11) | 0.0027 | 0.0334 | Xenobiotics | Food Component/Plant |
| ornithine | 0.42 | (0.14, 0.69) | 0.0028 | 0.0338 | Amino Acid | Urea cycle; Arginine and Proline Metabolism |
| X-24431 | -0.30 | (-0.50, -0.10) | 0.0028 | 0.0342 | N/A | N/A |
| glycochenodeoxycholate 3-sulfate | 0.34 | (0.12, 0.57) | 0.003 | 0.035 | Lipid | Primary Bile Acid Metabolism |
| gamma-glutamylisoleucine* | 0.37 | (0.13, 0.62) | 0.003 | 0.0353 | Peptide | Gamma-glutamyl Amino Acid |
| 5alpha-pregnan-3beta,20alpha-diol monosulfate (2) | -0.33 | (-0.55, -0.11) | 0.0031 | 0.0353 | Lipid | Progestin Steroids |
| 2-stearoyl-GPE (18:0)* | 0.34 | (0.12, 0.57) | 0.0032 | 0.036 | Lipid | Lysophospholipid |
| X-13658 | 0.33 | (0.11, 0.54) | 0.0033 | 0.036 | N/A | N/A |
| X-25420 | 0.45 | (0.15, 0.76) | 0.0033 | 0.036 | N/A | N/A |
| 2-hydroxy-3-methylvalerate | 0.36 | (0.12, 0.60) | 0.0034 | 0.037 | Amino Acid | Leucine, Isoleucine and Valine Metabolism |
| 1-carboxyethylphenylalanine | 0.35 | (0.12, 0.59) | 0.0035 | 0.037 | N/A | N/A |
| 5,6-dihydrouridine | 0.37 | (0.12, 0.62) | 0.0035 | 0.037 | Nucleotide | Pyrimidine Metabolism, Uracil containing |
| eicosapentaenoate (EPA; 20:5n3) | -0.33 | (-0.55, -0.11) | 0.0036 | 0.0375 | Lipid | Long Chain Polyunsaturated Fatty Acid (n3 and n6) |
| Fibrinopeptide B (1-9)** | 0.33 | (0.11, 0.55) | 0.004 | 0.0402 | Peptide | Fibrinogen Cleavage Peptide |
| sphingomyelin (d18:0/18:0, d19:0/17:0)* | 0.28 | (0.09, 0.46) | 0.004 | 0.0402 | Lipid | Dihydrosphingomyelins |
| X-12849 | -0.28 | (-0.47, -0.09) | 0.0041 | 0.0405 | N/A | N/A |
| 1-oleoyl-GPE (18:1) | 0.32 | (0.10, 0.54) | 0.0043 | 0.0424 | Lipid | Lysophospholipid |
| 4-hydroxyphenylpyruvate | 0.29 | (0.09, 0.49) | 0.0044 | 0.0425 | Amino Acid | Tyrosine Metabolism |
| glycochenodeoxycholate | 0.31 | (0.10, 0.52) | 0.0046 | 0.0437 | Lipid | Primary Bile Acid Metabolism |
| N1-Methyl-2-pyridone-5-carboxamide | -0.30 | (-0.50, -0.09) | 0.0047 | 0.044 | Cofactors and Vitamins | Nicotinate and Nicotinamide Metabolism |
| 2-hydroxy-4-(methylthio)butanoic acid | 0.33 | (0.10, 0.56) | 0.0047 | 0.044 | Amino Acid | Methionine, Cysteine, SAM and Taurine Metabolism |
| 7-alpha-hydroxy-3-oxo-4-cholestenoate (7-Hoca) | 0.43 | (0.13, 0.72) | 0.0052 | 0.0474 | Lipid | Sterol |
| X-11372 | -0.34 | (-0.57, -0.10) | 0.0052 | 0.0474 | N/A | N/A |
| palmitoleate (16:1n7) | 0.32 | (0.10, 0.55) | 0.0054 | 0.049 | Lipid | Long Chain Monounsaturated Fatty Acid |

**Table S6. Metabolites Associated with Anxiety Symptoms from Batch Combined Study Sample at Baseline *(FDR p-value < 0.05).*** Highlighted shaded metabolites indicate overlap with main findings (discovery-replication approach). Estimate indicates changes in STAI-10 (anxiety) continuous score per 1 SD increase in rank-normalized metabolite adjusting for age, sex, field center and Hispanic/Latino background. 95% CI represents 95% confidence intervals.

| **Metabolite** | **Estimate** | **95% CI** | **p** | **FDR p** | **Super Pathway** | **Sub Pathway** |
| --- | --- | --- | --- | --- | --- | --- |
| hydroxy-CMPF* | -0.63 | (-0.84, -0.41) | <0.0001 | <0.0001 | Lipid | Fatty Acid, Dicarboxylate |
| tyramine O-sulfate | 0.54 | (0.34, 0.74) | <0.0001 | <0.0001 | Amino Acid | Tyrosine Metabolism |
| 3-carboxy-4-methyl-5-propyl-2-furanpropanoate (CMPF) | -0.50 | (-0.71, -0.30) | <0.0001 | 0.0005 | Lipid | Fatty Acid, Dicarboxylate |
| beta-cryptoxanthin | -0.65 | (-0.91, -0.38) | <0.0001 | 0.0005 | Cofactors and Vitamins | Vitamin A Metabolism |
| X-24337 | 0.52 | (0.30, 0.74) | <0.0001 | 0.0009 | N/A | N/A |
| carotene diol (1) | -0.55 | (-0.80, -0.30) | <0.0001 | 0.0018 | Cofactors and Vitamins | Vitamin A Metabolism |
| 3-amino-2-piperidone | 0.61 | (0.33, 0.89) | <0.0001 | 0.0028 | Amino Acid | Urea cycle; Arginine and Proline Metabolism |
| X-24475 | -0.43 | (-0.64, -0.23) | <0.0001 | 0.0028 | N/A | N/A |
| X-11315 | -0.44 | (-0.65, -0.24) | <0.0001 | 0.0028 | N/A | N/A |
| 2-aminoheptanoate | 0.44 | (0.23, 0.66) | 0.0001 | 0.0057 | Lipid | Fatty Acid, Amino |
| X-24431 | -0.38 | (-0.58, -0.19) | 0.0001 | 0.0093 | N/A | N/A |
| threonate | -0.39 | (-0.59, -0.19) | 0.0002 | 0.0097 | Cofactors and Vitamins | Ascorbate and Aldarate Metabolism |
| docosahexaenoate (DHA; 22:6n3) | -0.47 | (-0.71, -0.22) | 0.0002 | 0.01 | Lipid | Long Chain Polyunsaturated Fatty Acid (n3 and n6) |
| 2-piperidinone | 0.46 | (0.22, 0.70) | 0.0002 | 0.0102 | Xenobiotics | Food Component/Plant |
| indolepropionate | -0.41 | (-0.63, -0.19) | 0.0002 | 0.0112 | Amino Acid | Tryptophan Metabolism |
| nicotinamide N-oxide | -0.38 | (-0.58, -0.18) | 0.0003 | 0.0125 | Cofactors and Vitamins | Nicotinate and Nicotinamide Metabolism |
| oxalate (ethanedioate) | -0.37 | (-0.57, -0.17) | 0.0003 | 0.0134 | Cofactors and Vitamins | Ascorbate and Aldarate Metabolism |
| alpha-tocopherol | -0.40 | (-0.63, -0.18) | 0.0004 | 0.0154 | Cofactors and Vitamins | Tocopherol Metabolism |
| gamma-tocopherol/beta-tocopherol | 0.44 | (0.20, 0.68) | 0.0004 | 0.0154 | Cofactors and Vitamins | Tocopherol Metabolism |
| 5alpha-pregnan-3beta,20alpha-diol monosulfate (2) | -0.42 | (-0.65, -0.18) | 0.0005 | 0.0194 | Lipid | Progestin Steroids |
| tartronate (hydroxymalonate) | -0.34 | (-0.54, -0.15) | 0.0006 | 0.0211 | Xenobiotics | Food Component/Plant |
| X-11372 | -0.42 | (-0.66, -0.18) | 0.0006 | 0.0214 | N/A | N/A |
| 1-(1-enyl-palmitoyl)-2-palmitoleoyl-GPC (P-16:0/16:1)* | 0.36 | (0.15, 0.57) | 0.0007 | 0.0237 | Lipid | Plasmalogen |
| X-24812 | 0.40 | (0.17, 0.63) | 0.0008 | 0.0246 | N/A | N/A |
| X-17010 | 0.54 | (0.23, 0.86) | 0.0008 | 0.0246 | N/A | N/A |
| 1-palmitoleoyl-GPC (16:1)* | 0.41 | (0.17, 0.65) | 0.0009 | 0.0246 | Lipid | Lysophospholipid |
| 1-arachidonoyl-GPE (20:4n6)* | 0.36 | (0.15, 0.57) | 0.0009 | 0.0246 | Lipid | Lysophospholipid |
| N-acetylputrescine | 0.39 | (0.16, 0.61) | 0.0009 | 0.0254 | Amino Acid | Polyamine Metabolism |
| 1-linoleoyl-GPE (18:2)* | 0.34 | (0.14, 0.55) | 0.0011 | 0.0281 | Lipid | Lysophospholipid |
| o-cresol sulfate | 0.41 | (0.16, 0.65) | 0.0011 | 0.0288 | Xenobiotics | Benzoate Metabolism |
| X-25419 | -0.48 | (-0.77, -0.19) | 0.0014 | 0.0342 | N/A | N/A |
| 3-phenylpropionate (hydrocinnamate) | -0.39 | (-0.64, -0.15) | 0.0016 | 0.0375 | Xenobiotics | Benzoate Metabolism |
| glucuronate | 0.37 | (0.14, 0.60) | 0.0016 | 0.0375 | Carbohydrate | Amino sugar Metabolism |
| methyl glucopyranoside (alpha + beta) | -0.32 | (-0.52, -0.12) | 0.0017 | 0.0375 | Xenobiotics | Food Component/Plant |
| N-methylproline | -0.33 | (-0.54, -0.12) | 0.0019 | 0.0418 | Amino Acid | Urea cycle; Arginine and Proline Metabolism |
| 1-methyl-4-imidazoleacetate | 0.41 | (0.15, 0.67) | 0.002 | 0.0418 | Amino Acid | Histidine Metabolism |
| 4-hydroxychlorothalonil | -0.34 | (-0.56, -0.13) | 0.0021 | 0.0428 | Xenobiotics | Chemical |
| choline | 0.35 | (0.13, 0.58) | 0.0022 | 0.0428 | Lipid | Phospholipid Metabolism |
| glutarate (C5-DC) | 0.35 | (0.13, 0.58) | 0.0022 | 0.0428 | Lipid | Fatty Acid, Dicarboxylate |
| X-11880 | -0.39 | (-0.63, -0.14) | 0.0024 | 0.0455 | N/A | N/A |
| X-25420 | 0.48 | (0.17, 0.79) | 0.0026 | 0.0455 | N/A | N/A |
| ethyl beta-glucopyranoside | -0.30 | (-0.50, -0.11) | 0.0026 | 0.0455 | Xenobiotics | Food Component/Plant |
| eicosapentaenoate (EPA; 20:5n3) | -0.32 | (-0.52, -0.11) | 0.0026 | 0.0455 | Lipid | Long Chain Polyunsaturated Fatty Acid (n3 and n6) |
| 2-hydroxy-4-(methylthio)butanoic acid | 0.35 | (0.12, 0.58) | 0.0026 | 0.0455 | Amino Acid | Methionine, Cysteine, SAM and Taurine Metabolism |
| 10-undecenoate (11:1n1) | -0.30 | (-0.50, -0.11) | 0.0027 | 0.0455 | Lipid | Medium Chain Fatty Acid |
| androstenediol (3beta,17beta) disulfate (1) | 0.36 | (0.13, 0.60) | 0.0027 | 0.0455 | Lipid | Androgenic Steroids |
| X-23593 | 0.30 | (0.10, 0.50) | 0.0029 | 0.0458 | N/A | N/A |
| ergothioneine | -0.41 | (-0.68, -0.14) | 0.003 | 0.0458 | Xenobiotics | Food Component/Plant |
| X-18901 | -0.32 | (-0.52, -0.11) | 0.003 | 0.0458 | N/A | N/A |
| 1-oleoyl-GPE (18:1) | 0.35 | (0.12, 0.58) | 0.003 | 0.0458 | Lipid | Lysophospholipid |

**Table S7. Associations between Identified Metabolites and Depression Symptoms from Longitudinal Mixed Effects Model (Batch 1 and 2, both visits).** Estimate indicates changes in CES-D-10 (depression) continuous score per 1 SD increase in rank-normalized metabolite adjusting for age, sex, field center and Hispanic/Latino background with a random intercept for individual ID and household ID. 95% CI represents 95% confidence intervals.

| **Metabolite** | **Estimate** | **95% CI** | **p** | **FDR p** |
| --- | --- | --- | --- | --- |
| beta-cryptoxanthin | -0.54 | (-0.71, -0.38) | 7.77E-11 | 1.94E-10 |
| tyramine O-sulfate | 0.45 | (0.31, 0.59) | 8.29E-10 | 1.38E-09 |
| hydroxy-CMPF* | -0.54 | (-0.69, -0.39) | 1.64E-12 | 8.21E-12 |
| choline | 0.36 | (0.21, 0.51) | 1.67E-06 | 1.67E-06 |
| X-24337 | 0.40 | (0.25, 0.55) | 1.83E-07 | 2.28E-07 |

**Table S8. Causal Effects of Selected Metabolites on Depression and Anxiety Symptoms from One-Sample Mendelian Randomization.**

| **GWAS Study for PRS** | **Number of SNPs** | **SNPs** | **Exposure (Metabolite)** | **Outcome** | **Beta** | **SE** | ***P*-value** |
| --- | --- | --- | --- | --- | --- | --- | --- |
| Yin, X., et al. (2022). Genome-wide association studies of metabolites in Finnish men identify disease-relevant loci. Nature Communications, 13(1), 1644. | 2 | rs149133795, rs2807074 | 3-amino-2-piperidone | anxiety (STAI-10) | 3.89E+00 | 4.56E+01 | 9.32E-01 |
|  | 2 | rs566530, rs11188161 | hydroxy-CMPF | depression (CES-D-10) | 1.98E+00 | 1.55E+01 | 8.98E-01 |
|  | 2 | rs566530, rs11188161 | hydroxy-CMPF | anxiety (STAI-10) | 2.66E+00 | 1.70E+01 | 8.76E-01 |
|  | 9 | rs2392848, rs10846744, rs2970082, rs117105192, rs12924312, rs12103224, rs11643509, rs34798987, rs146627417 | carotene-diol-1 | anxiety (STAI-10) | 5.24E-01 | 4.62E+00 | 9.10E-01 |
|  | 10 | rs12524503, rs4646352, rs113707589, rs141107190, rs55715619, rs72802356, rs75925066, rs6870496, rs72788669, rs115210424 | choline | depression (CES-D-10) | -4.15E-01 | 1.62E+01 | 9.80E-01 |
